# Driving novel endpoints and study designs in amyotrophic lateral sclerosis: closer examination of the ALSFRS-R subdomains and a new definition of fast and slow progressors

**DOI:** 10.1101/2025.08.21.25334108

**Authors:** Robert Ellis, William B. Nowell, Nikesh Patel, Matthew Wipperman, Jiangnan Lyu, Shawn Mishra, Anthony Scotina, Danni Tu, John A. Wagner, Oren Levy, The Pooled Resource Open-Access ALS Clinial Trials Consortium

## Abstract

Amyotrophic lateral sclerosis (ALS) is a motor neuron disease that leads to significant morbidity and mortality, but with substantial variability in the rate of progression [1,2]. A common primary outcome measure used in ALS clinical studies is the ALSFRS-R. Change in this measure may also be used to stratify fast and slow progressors, typically with the assumption of linear decline. Reported limitations of the ALSFRS-R and the assumption of linear slope may hinder novel therapy development. Here we use two distinct populations from a clinical trial database (PRO-ACT) and a natural history registry (ALS TDI) to show that ALSFRS-R fine and gross motor subdomains are more sensitive to disease progression than total score and bulbar and respiratory subdomain scores. We also present a novel non-linear method for defining fast and slow progressors. Together these results may improve demonstration of outcomes in ALS interventional studies using the ALSFRS-R.

## Introduction

Since the approval of edaravone in 2017, drug development in amyotrophic lateral sclerosis (ALS) has encountered a number of failed trials for novel therapies (e.g. NCT05237284, NCT05866926, NCT04944784, NCT05842941, NCT05740813). For decades, the clinician-reported ALS Functional Rating Scale - Revised (ALSFRS-R) [3] has been the gold standard in ALS clinical trial measurement; trials evaluating therapies for ALS are typically configured with a duration of 6 to 12 months and a primary endpoint of change from baseline in ALSFRS-R total score. Furthermore, the ALSFRS-R is the only clinical outcome assessment appearing in ALS drug labels [4] and is recommended for ALS drug development in both EMA [5] and FDA guidelines [6]. Despite its widespread use, concept validity, and demonstrated reliability [7,8], the ALSFRS-R has several reported limitations: (1) the total score lacks sensitivity to early changes in disease, particularly among patients whose ALS is progressing slowly; (2) items and subdomains are equally weighted despite uneven subdomain response [9], uneven item structure [10], and low probability of observation for some item scores [11]; and, (3) an assumption of a linear decline in function despite observed between-visit variability of up to 5 points [12]. These limitations of the ALSFRS-R, along with significant heterogeneity in disease progression over time both within and between ALS patients, present challenges to accurately assessing treatment effect in interventional clinical trials.

The estimated decline in ALSFRS-R total score over a specific time period has been used to support stratification of a study population into fast and slow progressors [13,14,15]. Characterization of linear functional decline has been used in trial inclusion and stratification criteria in an effort to more efficiently detect treatment effect in clinical trials (e.g. NCT02623699). Accumulating evidence suggests ALS progression is likely to be non-linear (Supplemental Figure 1) [16,17,18,19]. If indeed ALS progression is non-linear, then the use of linear methods to estimate decline in ALSFRS-R total score risks mischaracterizing change over time, and may impair the detection of treatment effects. While different analyses have demonstrated that progression is likely to be non-linear, a deterministic method for estimating non-linear progression that can be used for subject stratification in a prospective study has yet to be defined.

Exploratory retrospective longitudinal analysis of ALSFRS-R scores was conducted to address two main questions: 1) are subdomain scores of the ALSFRS-R more sensitive to disease progression than the ALSFRS-R total score? and, 2) can deterministic non-linear methods be developed to more accurately characterize ALS progression to support subject stratification using ALSFRS-R total score? Answers to these questions may enhance researchers’ use of information provided by the ALSFRS-R to more accurately monitor disease progression and assess treatment impact.

## Methods

### Study design and data source

Datasets used for all analyses were obtained from Pooled Resource Open-Access ALS Clinical Trials (PRO-ACT) and ALS Therapy Development Institute (ALS TDI). The data available in the PRO-ACT Database have been volunteered by PRO-ACT Consortium members and contains aggregated ALS patient records collected between 1990 and 2010 from 16 existing publicly- and privately-conducted clinical trials and one observational study [20]. PRO-ACT was created by the non-profit organization Prize4Life in partnership with the Northeast ALS Consortium, the Neurological Clinical Research Institute at Massachusetts General Hospital, and with funding from the ALS Therapy Alliance. The ALS Research Collaborative (ARC) of the ALS TDI is an ongoing observational study in which patients provide data directly and remotely (ALS Therapy Development Institute, Watertown, MA, USA). The dataset was downloaded from the ARC Data Commons and includes patient-reported ALSFRS-R data from 2014 to 2024 for 1,426 patients voluntarily participating in the natural history registry and who may have concurrently participated in an interventional study. Both datasets were licensed under data agreements from their respective owners and contained de-identified data. Ethical approval for PRO-ACT and ALS TDI data collection was granted from their respective Institutional Review Boards.

PRO-ACT ALSFRS-R scores are clinician-reported and begin at baseline and conclude at withdrawal from study or patient death. ALS TDI ALSFRS-R scores are self-reported and begin at patient registration and conclude at withdrawal from registry, patient death, or end of available data. While there is high correlation between ALSFRS-R scores reported by clinicians and by patients, patients tend to score themselves slightly higher than clinicians, with the limits of agreement between clinician and self-reported ALSFRS-R estimated to be 4.3 points [21].

Separate analytic datasets were created from the PRO-ACT and ALS TDI datasets, and these analytic datasets included subjects with baseline disease history, demographic data (e.g. age and sex), and per visit ALSFRS-R scores for at least 24 weeks with a minimum of five visits. Analyses were limited to the first 24 weeks of data except for the ALS TDI mock study. A duration of 24 weeks was chosen because it is routinely used in ALS interventional studies [22] and has approval precedent [23]. Summary statistics and histograms were plotted for all variables in both analytic datasets to evaluate covariate distributions. Analyses of both datasets were completed with Python 3.9 (Python Software Foundation) with numpy v1.25, scipy v1.13.1 and semopy v2.3.11.

### ALSFRS-R total score and subdomain scores

The ALSFRS-R is an ordinal scale of 12 items grouped into four subdomains of three items each: bulbar, fine motor, gross motor, and respiratory. Individual items are scored 4 (normal) to 0 (severe), reflecting a patient’s ability or need for assistance with certain activities and functions; ratings for each item within a subdomain are summed to a subdomain score (range 0 to 12), and subdomain scores are summed to a total score (range 0 to 48) [24]. The measure is alternatively characterized as having three subdomains when fine and gross motor are combined into a single “motor” subdomain of six items [10]. `The minimum clinically important difference (MCID) for decline/worsening is estimated to be ≥ 4 points` [25].

Confirmatory factor analysis (CFA) was performed on week zero ALSFRS-R scores for placebo-treated subjects in the PRO-ACT dataset and registry subjects in the ALS TDI dataset. The analysis was conducted to verify the measure’s subdomain structure before proceeding with responsiveness analyses to evaluate the relative sensitivity of subdomain scores to ALS disease progression as compared with that of the ALSFRS-R total score. Goodness of fit of one-, three-, and four-factor models, as well as bi-factor models with three- and four factors, were evaluated.

Item score distributions by actual score and score probability were examined to determine whether the ALSFRS-R total score and each of the subdomain scores similarly and evenly capture disease progression. Score probability was defined as the number of instances of a score over the total number of observed scores by subdomain. Score distribution skew from normal was calculated.

### Characterization of ALS disease progression

Longitudinal progression characteristics were evaluated by calculating the 7-day rolling mean and weekly quartiles of change from baseline in ALSFRS-R total and subdomain scores. Rolling mean change was chosen because the PRO-ACT dataset represents the aggregate of multiple different study visit schedules. A period of seven days was chosen to analyze weekly change. Over a 24-week period, change by week from baseline in ALSFRS-R total and subdomain scores was examined as a percentage of maximum score and in absolute terms to determine whether specific subdomains may be more sensitive measures of disease progression compared to ALSFRS-R total score.

### Development of a non-linear progression model

To estimate non-linear disease progression prospectively, for the purpose of cohort stratification, an adaptation of the finite difference method [26] was implemented.

The progression model developed in this analysis uses slope between each visit instead of an aggregate slope over all visits. To develop the model, subjects in each dataset were required to have more than the median number of available ALSFRS-R scores between baseline and 24 weeks. Between-visit slope was calculated using the ALSFRS-R total score and the resulting slope was classified as positive, negative, or no change (Supplemental Figure 2). The percentages of time on study spent in each slope classification were used to define thresholds for four possible progression cohorts that have been reported in the literature: fast, slow, no change and alternating change. Alternating change corresponds with the reported observation that some subjects show periods of both functional improvement (positive slope) and decline (negative slope) [13]. Spaghetti plots were generated for each progression cohort to allow for visual comparison of change in ALSFRS-R total score from baseline in each cohort. Four different analyses were performed to evaluate non-linear model utility: 1) comparison of cohort membership by linear and non-linear progression models; 2) sensitivity analysis for number of visits required to classify patient cohort; 3) change in ALSFRS-R subdomain scores between linear and non-linear models; and, 4) a mock study using the ALS TDI dataset. To estimate the minimum number of visits required for the non-linear model to classify all subjects, the number of classified and unclassified subjects was evaluated as a function of the number of visits used in the model.

Linear progression was defined as average slope for available measures. This was computed to support comparison between linear and non-linear progression models. The linear model classifies patients as negative or positive slope, or as unclassified. Both linear and non-linear progressor classification was calculated for all subjects in both datasets with more than five visits.

A mock study was constructed from the ALS TDI dataset to simulate an interventional study design in which retrospective ALSFRS-R data are used to: 1) stratify patients at baseline into fast and slow progression cohorts, and 2) examine prospective change forward from baseline in the same cohorts. ALS TDI data over 48 weeks were split into two halves: 24 weeks of “retrospective” and 24 weeks of “prospective” data. Subjects with five or more visits in the retrospective data were identified and the non-linear model was applied to their data. Fast progressing subjects were classified using the thresholds defined in model development (Table 2). All other subjects in the retrospective data were assigned to the slow progressing cohort. Mean change from baseline in ALSFRS-R total and subdomain scores for the retrospective period were then calculated for the fast and slow progressing prospective cohorts. The non-linear model was then applied to the mock 24-week prospective data and the same scores calculated. The specificity, sensitivity, and accuracy of the retrospective cohort allocation were calculated by counting the number of prospective subjects identified as fast and slow progressors retrospectively versus classified prospectively. Lastly, the linear model was applied to the same retrospective and prospective datasets, and classification of progression cohort by non-linear and linear models was compared.

## Results

### Subjects in the PRO-ACT clinical trial dataset have more recent diagnosis and are less likely to have definite ALS diagnosis than subjects in the ALS TDI registry dataset

PRO-ACT and ALS TDI datasets were similar in distribution of mean and median baseline ALSFRS-R total score, subject gender, and subject age at enrollment (Table 1, Supplemental Figure 3). Differences were identified between the two datasets for mean (sd) of weeks of study duration (PRO-ACT, 41.5 (19.0); ALS TDI, 81.9 (109.3)) and number of completed ALSFRS-R responses per subject (PRO-ACT, 9.3(4.3); ALS TDI 29.6 (91.3)). The PRO-ACT dataset was composed of subjects who were more recently diagnosed (t-test, p-value < 0.0001) and less likely to have an El Escorial definite ALS diagnosis [27] in dataset disease history (chi-square, p-value < 0.0001) than in the ALS TDI dataset.

**Table 1.**
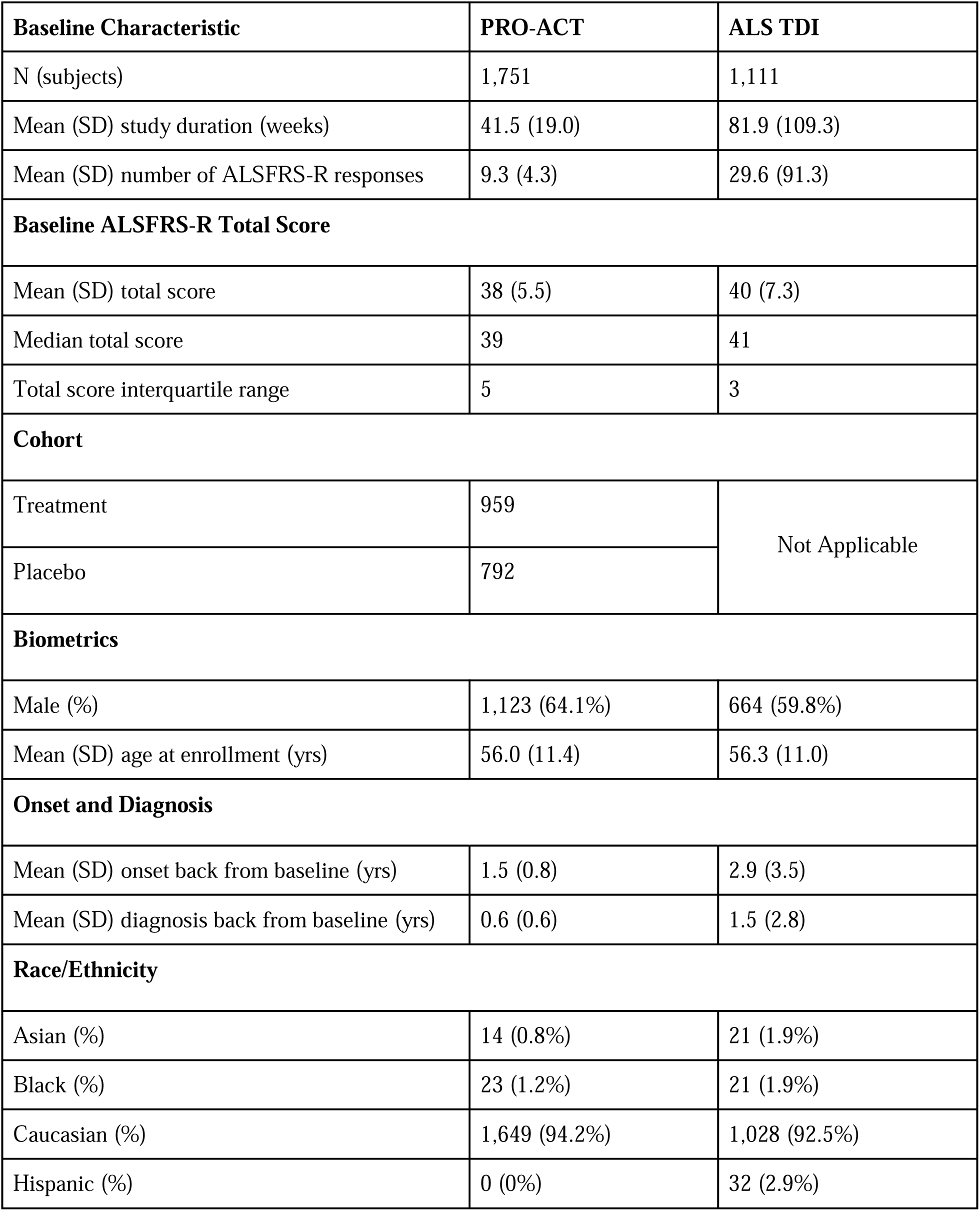
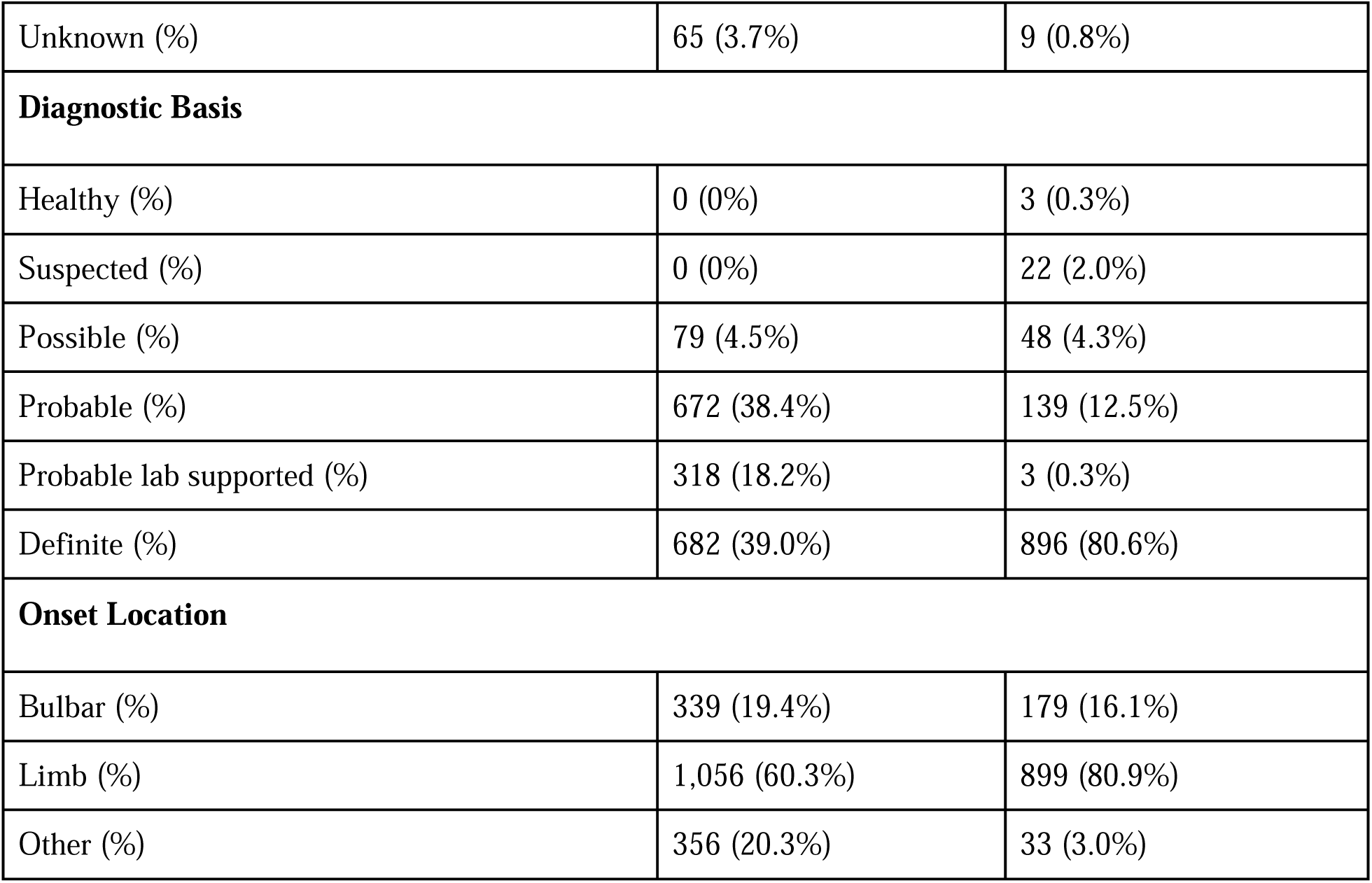
Baseline disease history and biometric characteristics of the PRO-ACT (clinical trial) and ALS TDI (natural history registry) analysis datasets.

| Baseline Characteristic | PRO-ACT | ALS TDI |
| --- | --- | --- |
| N (subjects) | 1,751 | 1,111 |
| Mean (SD) study duration (weeks) | 41.5 (19.0) | 81.9 (109.3) |
| Mean (SD) number of ALSFRS-R responses | 9.3 (4.3) | 29.6 (91.3) |
| Baseline ALSFRS-R Total Score |  |  |
| Mean (SD) total score | 38 (5.5) | 40 (7.3) |
| Median total score | 39 | 41 |
| Total score interquartile range | 5 | 3 |
| Cohort |  |  |
| Treatment | 959 | Not Applicable |
| Placebo | 792 |  |
| Biometrics |  |  |
| Male (%) | 1,123 (64.1%) | 664 (59.8%) |
| Mean (SD) age at enrollment (yrs) | 56.0 (11.4) | 56.3 (11.0) |
| Onset and Diagnosis |  |  |
| Mean (SD) onset back from baseline (yrs) | 1.5 (0.8) | 2.9 (3.5) |
| Mean (SD) diagnosis back from baseline (yrs) | 0.6 (0.6) | 1.5 (2.8) |
| Race/Ethnicity |  |  |
| Asian (%) | 14 (0.8%) | 21 (1.9%) |
| Black (%) | 23 (1.2%) | 21 (1.9%) |
| Caucasian (%) | 1,649 (94.2%) | 1,028 (92.5%) |
| Hispanic (%) | 0 (0%) | 32 (2.9%) |
| Unknown (%) | 65 (3.7%) | 9 (0.8%) |
| **Diagnostic Basis** |  |  |
| Healthy (%) | 0 (0%) | 3 (0.3%) |
| Suspected (%) | 0 (0%) | 22 (2.0%) |
| Possible (%) | 79 (4.5%) | 48 (4.3%) |
| Probable (%) | 672 (38.4%) | 139 (12.5%) |
| Probable lab supported (%) | 318 (18.2%) | 3 (0.3%) |
| Definite (%) | 682 (39.0%) | 896 (80.6%) |
| **Onset Location** |  |  |
| Bulbar (%) | 339 (19.4%) | 179 (16.1%) |
| Limb (%) | 1,056 (60.3%) | 899 (80.9%) |
| Other (%) | 356 (20.3%) | 33 (3.0%) |

### The range of change in ALSFRS-R total score from baseline at 24 weeks is greater than fifty percent of the possible score range

At 24 weeks, the mean change in ALSFRS-R total score differed between the two datasets (PRO-ACT, ∼35 points; ALS TDI, ∼25 points), and quartile ranges of 35 and 25 points, respectively, represent 72% and 52% of the ALSFRS-R potential total score change of 48 points. Furthermore, in both datasets, the quartile range at 24 weeks was ≥ 6 times the estimated MCID threshold for decline of ≥ 4 points [25].

### Fine and gross motor subdomain scores are sensitive measures of disease progression at 24 weeks

#### Confirmatory factor analysis

According to the CFA fit indices, the bifactor model with four subdomain factors and a fifth general factor (representing ALSFRS-R total score) provided the best fit for the data (Supplemental Table 1). The chi-square value was significant, [PRO-ACT: □^2^ (df 32, N=792) = 85.302, p<0.001; ALS TDI: □^2^ (df 32, N=1,111) = 139.547, p<0.001], indicating poor fit. However, chi-square tests are sensitive to sample size; when sample size is large, the test will often indicate significance. The alternative goodness of fit statistics indicated an acceptable fit between model and data: [PRO-ACT: RMSEA 0.052, TLI 0.968, CFI 0.984; ALS TDI: RMSEA 0.049, TLI 0.980, CFI 0.990]. Comparing the nested models yielded a statistically significant difference between the four-factor + general factor model and the three-factor + general factor model [PRO-ACT: Δ □^2^ (Δdf=4, N=792) = 28.644, p<0.001; ALS TDI: Δ □^2^ (Δdf=4, N=1,111) = 748.319]. Therefore, the three-factor + one-factor model does not fit as well as the four-factor + general model, providing evidence that the ALSFRS-R reflects overall ALS patient function with four functional subdomains: bulbar, fine motor, gross motor, and respiratory.

#### Comparison of ALSFRS-R total and subdomains score distribution and change

The distribution of ALSFRS-R total score in both datasets was similar with 50% cumulative probability of total score > 36 for PRO-ACT and > 39 for ALS TDI (Figures 2a and 2b). The probability of severe impairment subdomain scores (< 4 points) was < 10% in both PRO-ACT and ALS TDI populations, and normal scores (12 points) were more frequently observed for respiratory and bulbar subdomains than for fine motor and gross motor subdomains (Figures 2c and 2d, Supplemental Figure 4). The fine and gross motor subdomains in both datasets were moderately skewed (skew between zero and 0.5). The bulbar and respiratory subdomains in both datasets were highly skewed (skew > 2), indicating that the analyzed study populations did not include a large number of participants with severe respiratory and/or bulbar impairments. Mean decline in ALSFRS-R total scores was greater overall in the PRO-ACT dataset (mean % change [CI]: 11.7% [−13.5%,−10.0%]) than in the ALS TDI dataset (mean % change [CI]: −6.3% [−7.2%,−5.4%]) (Supplemental Table 2, Figures 3a and 3b).

**Figure 1.**
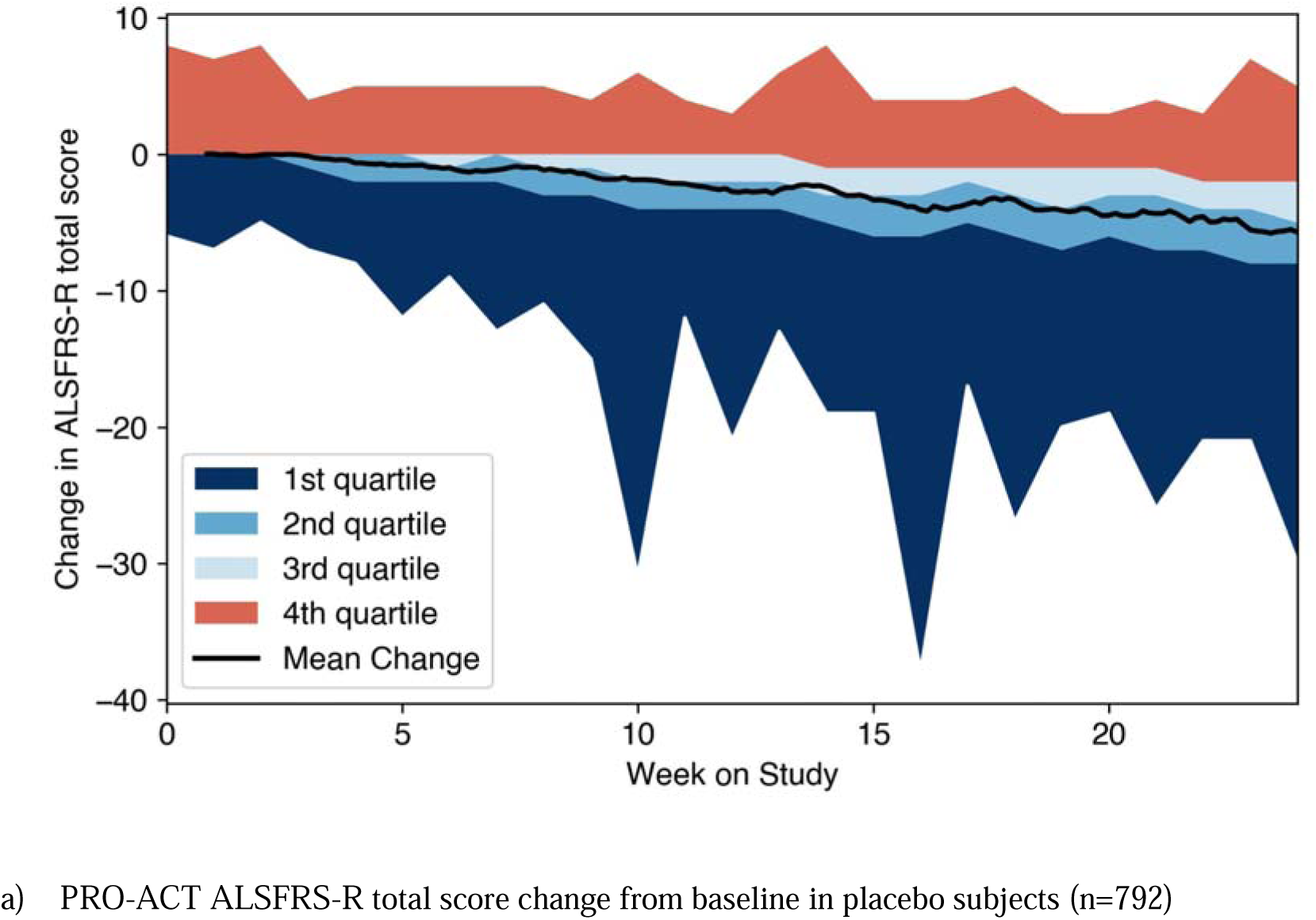
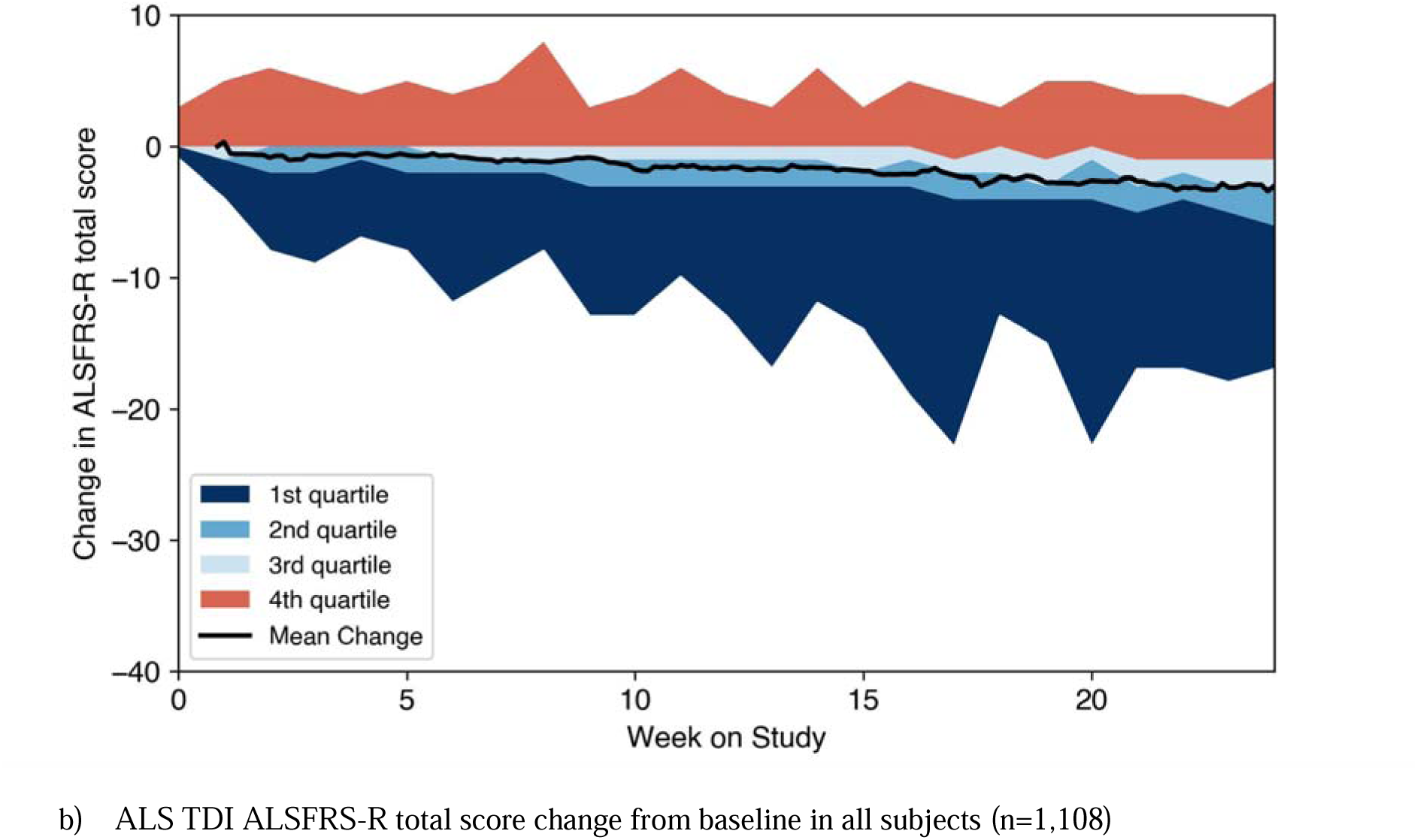
ALSFRS-R change from baseline in the PRO-ACT and ALS TDI datasets. Plots show the rolling 7-day change in ALSFRS-R total score for all subjects in each dataset between weeks 0 (baseline) and 24. Shaded areas represent change from baseline quartile with 1st quartile representing the largest change and 4th quartile the least change from baseline. In both datasets a small number (<5) of subjects only have data before baseline and are not plotted.

**Figure 2.**
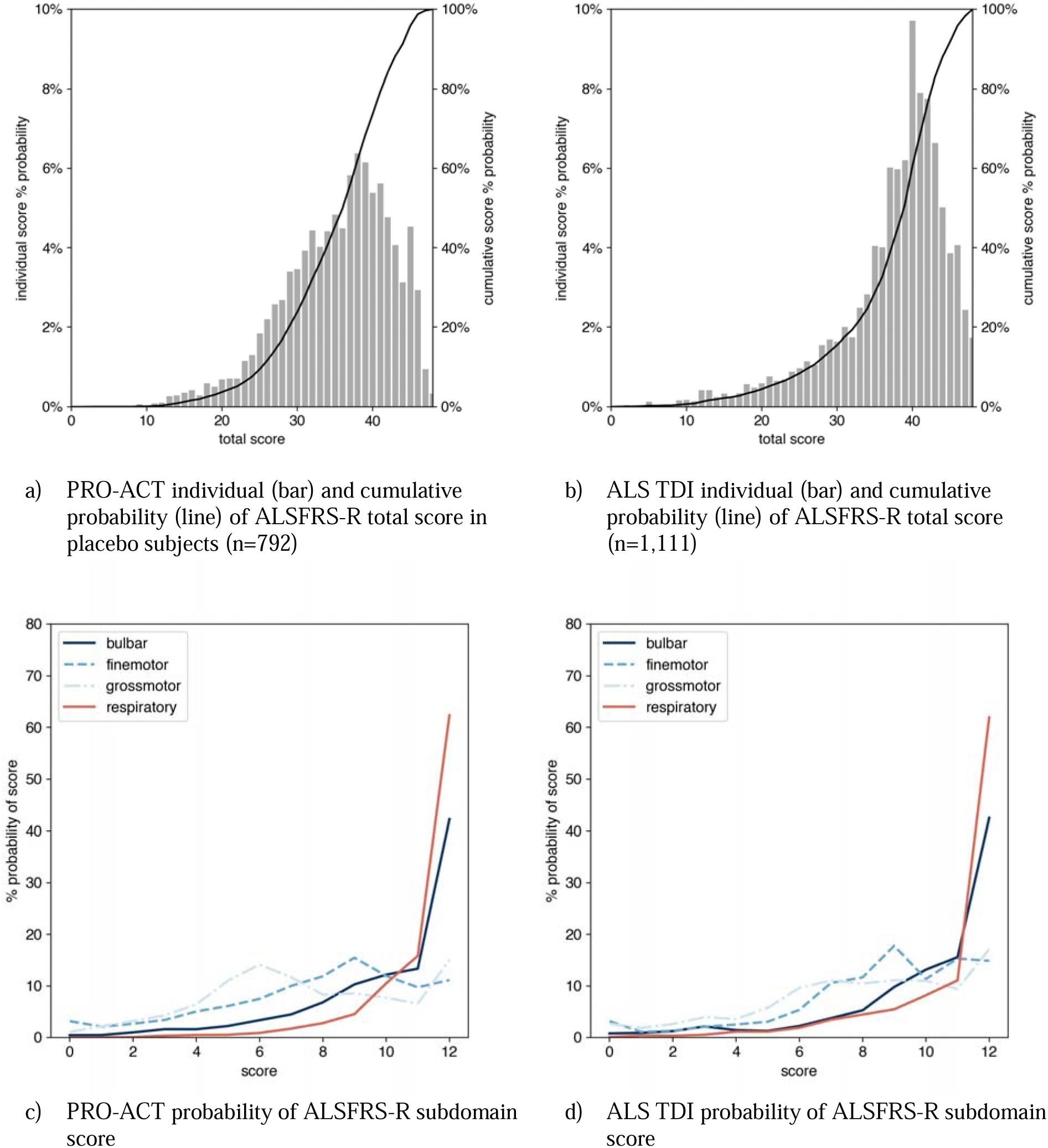
ALSFRS-R total and subdomain scores and probabilities. In the PRO-ACT (subplots a and c) and ALS TDI (subplots b and d) datasets over 24 weeks for all patients: 1,751 for PRO-ACT and 1,111 for ALS TDI. Total score range is zero to 48 and each subdomain score range is zero to 12.

**Figure 3.**
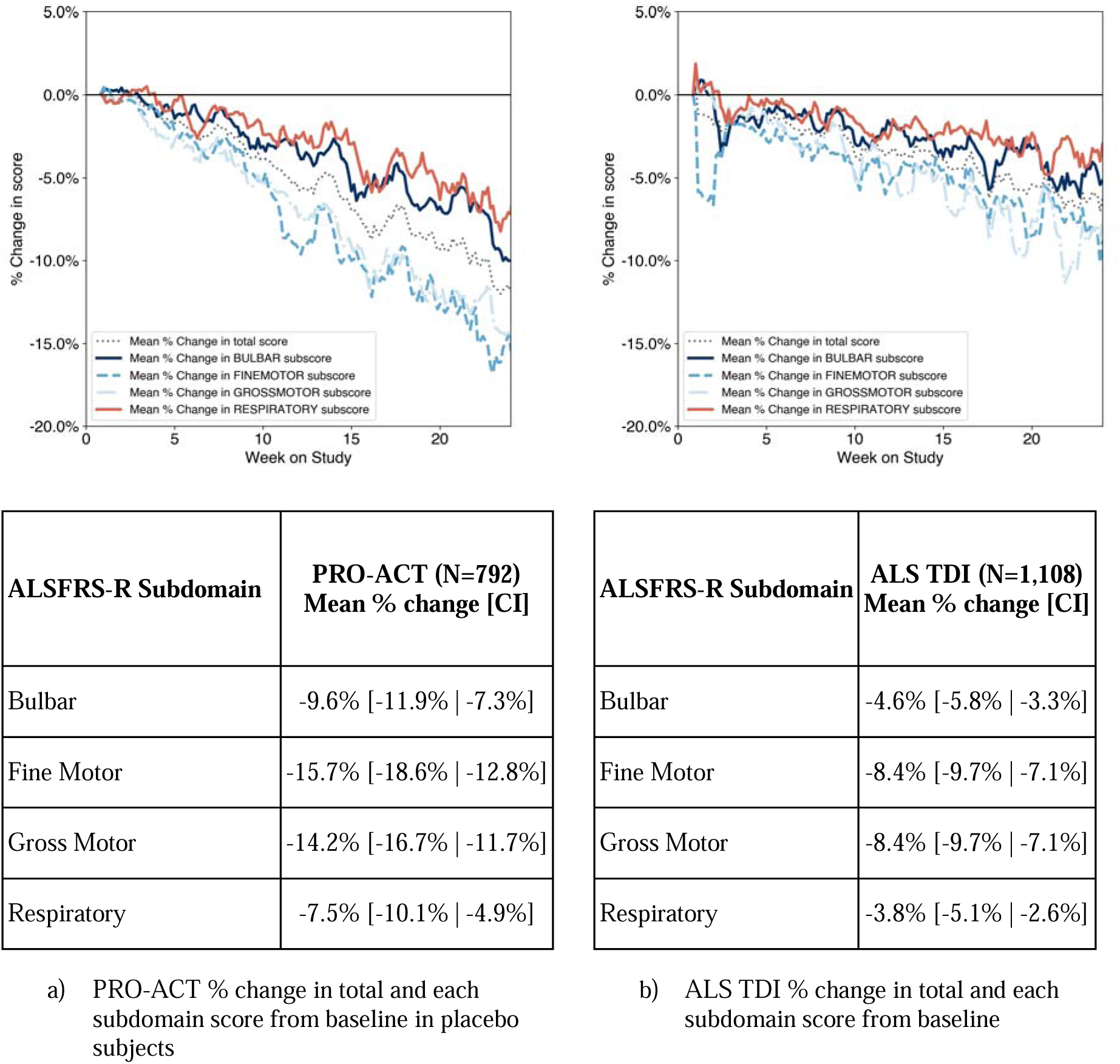
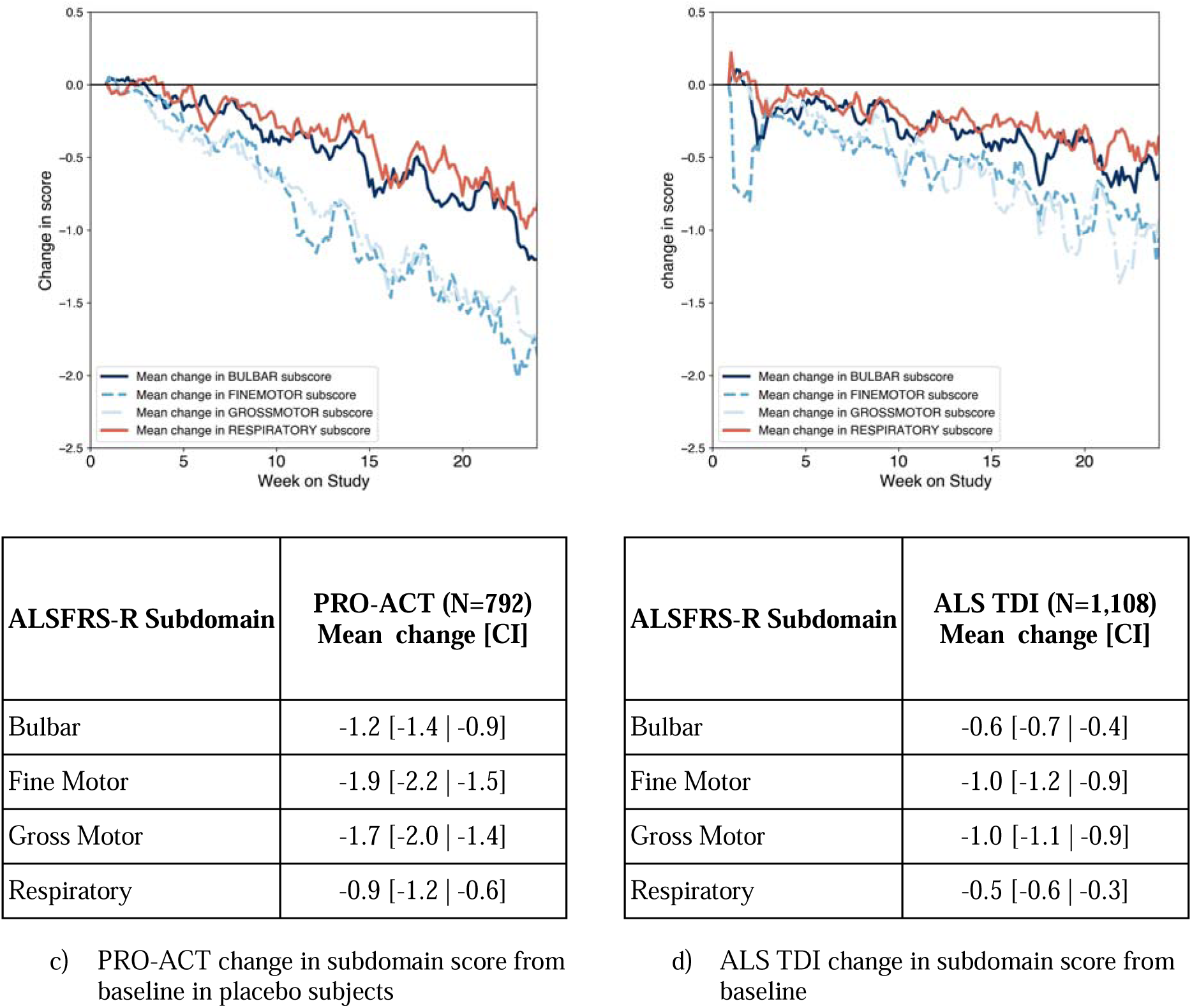
Percent change in total and subdomain score and actual subdomain score change from baseline to week 24 for PRO-ACT and ALS TDI datasets. Subplots a) and c) show PRO-ACT percent change for total and subdomain scores then actual change in subdomain score in placebo subjects (n=792). Subplots b) and d) show the same for the ALS TDI dataset (N=1,111).

Fine and gross motor subdomain scores showed greater percent change at 24 weeks (mean [CI]: PRO-ACT, −14 to −16% [−19%,−12%]; ALS TDI −8% [−10%, −7%]), than total score (mean [CI]:PRO-ACT, −12% [−14%,−10%]; ALS TDI −6% [−7%,−5%]) or other subdomain scores (Figure 3, Supplemental Table 2). Both datasets maintained an approximate order of fastest to slowest progressing score: fine motor / gross motor subdomain score, total ALSFRS-R score, bulbar subdomain score, and respiratory subdomain score.

### Non-linear progression can be quantified and used to stratify patients with ALS

Contour plots of slope category distribution showed similar patterns across both datasets (Supplemental Figure 5), with some subjects spending up to 80% of time on study with negative between-visit slope (decline) and other subjects split between negative, positive and no change between-visit slope.

Visual inspection of the line plot of percent of subjects by percent time in study for each type of slope category (Supplemental Figure 6) suggested that the proportion of subjects by slope type splits before and after 40-50% time spent on study. Specifically, subjects were more likely to have positive or no slope before 40-50% time spent on study, and more likely to have no change or negative slope after 40-50% time spent on study. Spaghetti plots of the four candidate progression cohorts (fast, slow, alternating, and no change using thresholds in Table 2) revealed clear separation between the fast and no progression cohorts in both datasets (Figures 4a to 4d). Slow and alternating progression cohort plots showed similar patterns of variability and different total decline, indicating that those in the alternating cohort experienced greater decline over time than those in the slow cohort (Figures 4e to 4h).

**Table 2.** Proof of concept thresholds for 4 progression cohort definitions. Thresholds are defined using percent of time on study in each of negative, positive or no change in slope. Note that the 4-cohort definition below means that some subjects may be unclassified.

| Progression Cohort | Threshold Definition |
| --- | --- |
| Fast progressor | > 50% of time with negative change/decline in ALSFRS-R total score |
| Not progressing | > 50% of time with no change in ALSFRS-R total score |
| Slow progressor | > 40% of time with positive change in ALSFRS-R total score |
| Alternating progressor | > 25% of time with positive and > 25% of time with negative change in ALSFRS-R total score and not already classified as fast or slow |

**Figure 4.**
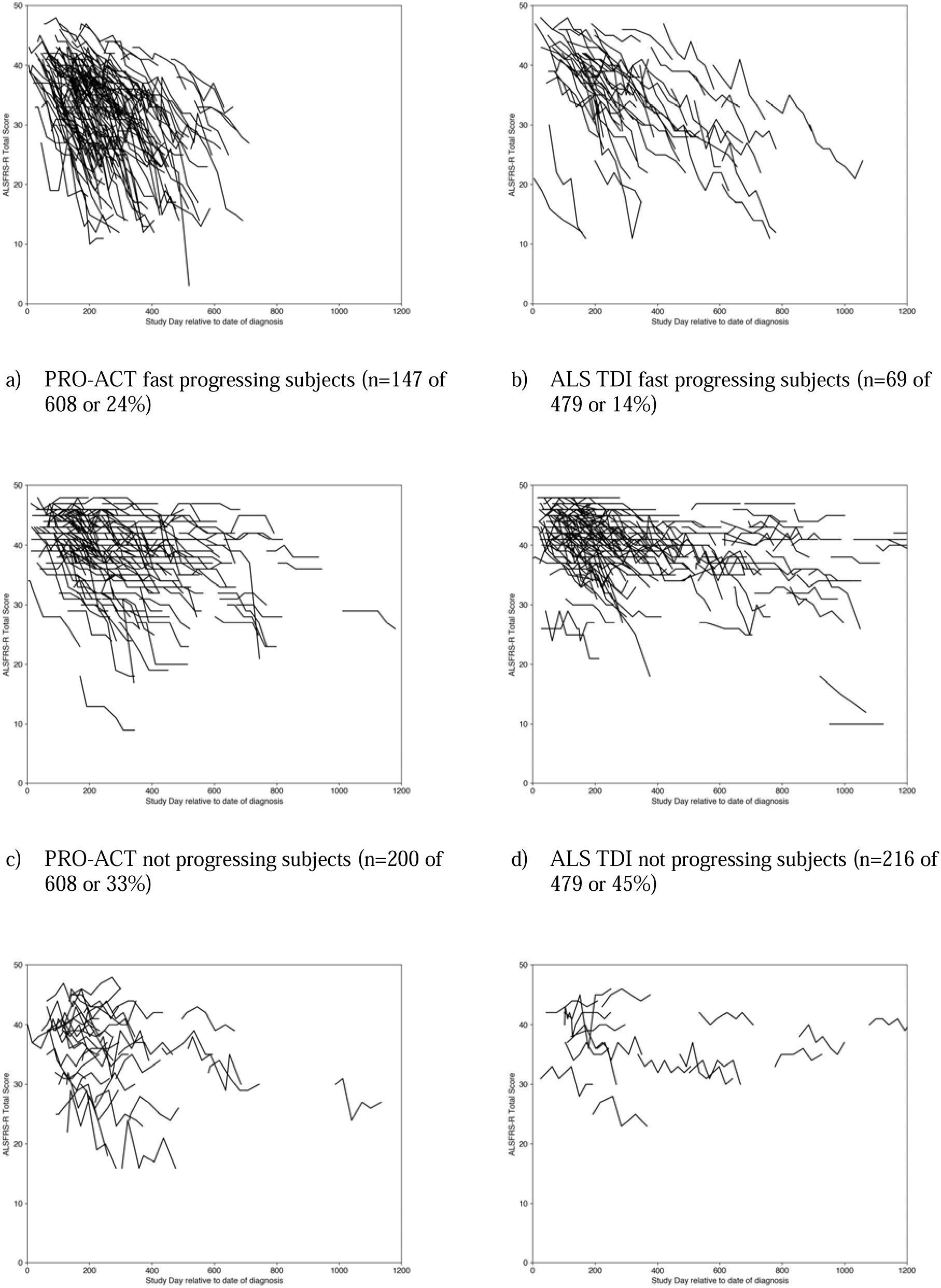
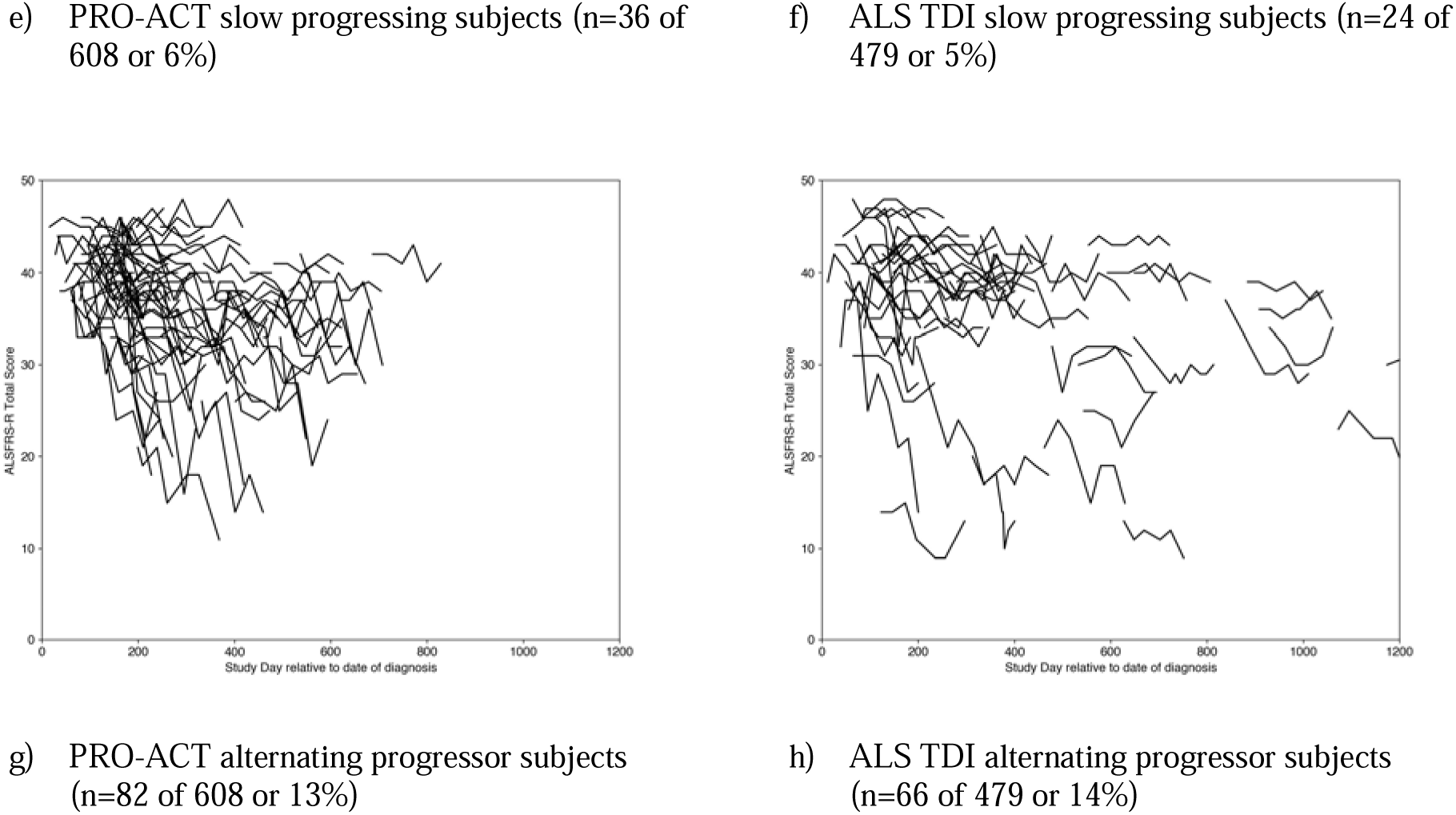
Spaghetti plots of subject ALSFRS-R total score by progression category plotted relative to diagnosis to aid visual separation of each subject. Subplots a, c, e and g show results for the PRO-ACT dataset and subplots b, d, f and h for the ALS TDI dataset.

Comparing linear and non-linear methods of progression classification, the linear model classified > 80% of subjects as having negative slope and the non-linear model classified > 95% of subjects as having mixed slope (Table 3). The linear model cannot, by design, account for variability in slope over time and therefore estimates a mean overall rate of decline.

**Table 3.**
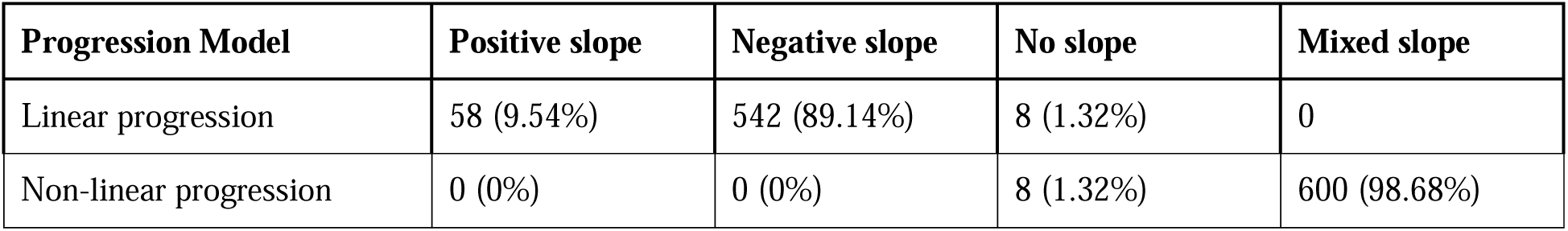
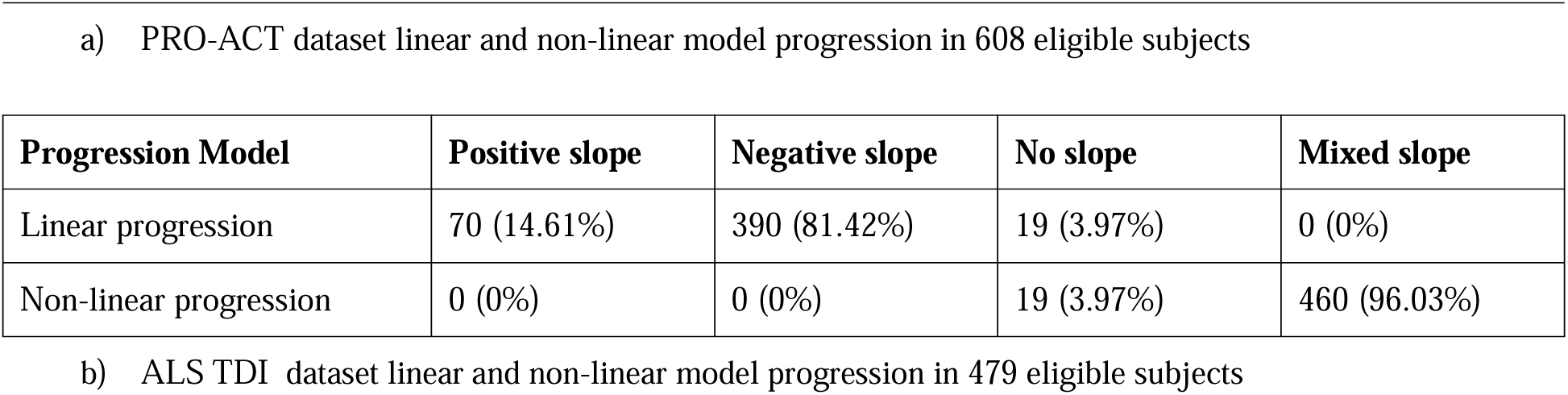
Comparison of linear and non-linear progression models in a) PRO-ACT and b) ALS TDI datasets. Subjects with > 5 visits between baseline and 24 weeks were classified as positive, negative, no or mixed slope. Mixed slope is defined as any combination of more than one type of slope. Alternating progressors are a subset of mixed slope. Table cells show the count of subjects and percent of total subjects.

With a minimum of five visits, the four-cohort definition classified 76% of PRO-ACT (n=608) and 78% of ALS TDI (n=479) subjects with ∼25% remaining unclassified. With a minimum of three visits as the requirement, the proportion of unclassified subjects increased to ∼33%. This suggests that a minimum of five visits is preferable for successful progression cohort stratification with this model.

### Non-linear stratified fast and slow progressors in a mock study

The ALS TDI dataset was used to construct a mock study in which ALSFRS-R scores prior to baseline are used to stratify patients as fast or slow progressors for prospective study (Figure 5a). A total of 548 of 667 evaluable subjects in the mock retrospective study data met the requirement of five or more visits in weeks 0 to 24. Of these 548 subjects, 488 (89%) also had ALSFRS-R scores in the “prospective” dataset.

**Figure 5.**
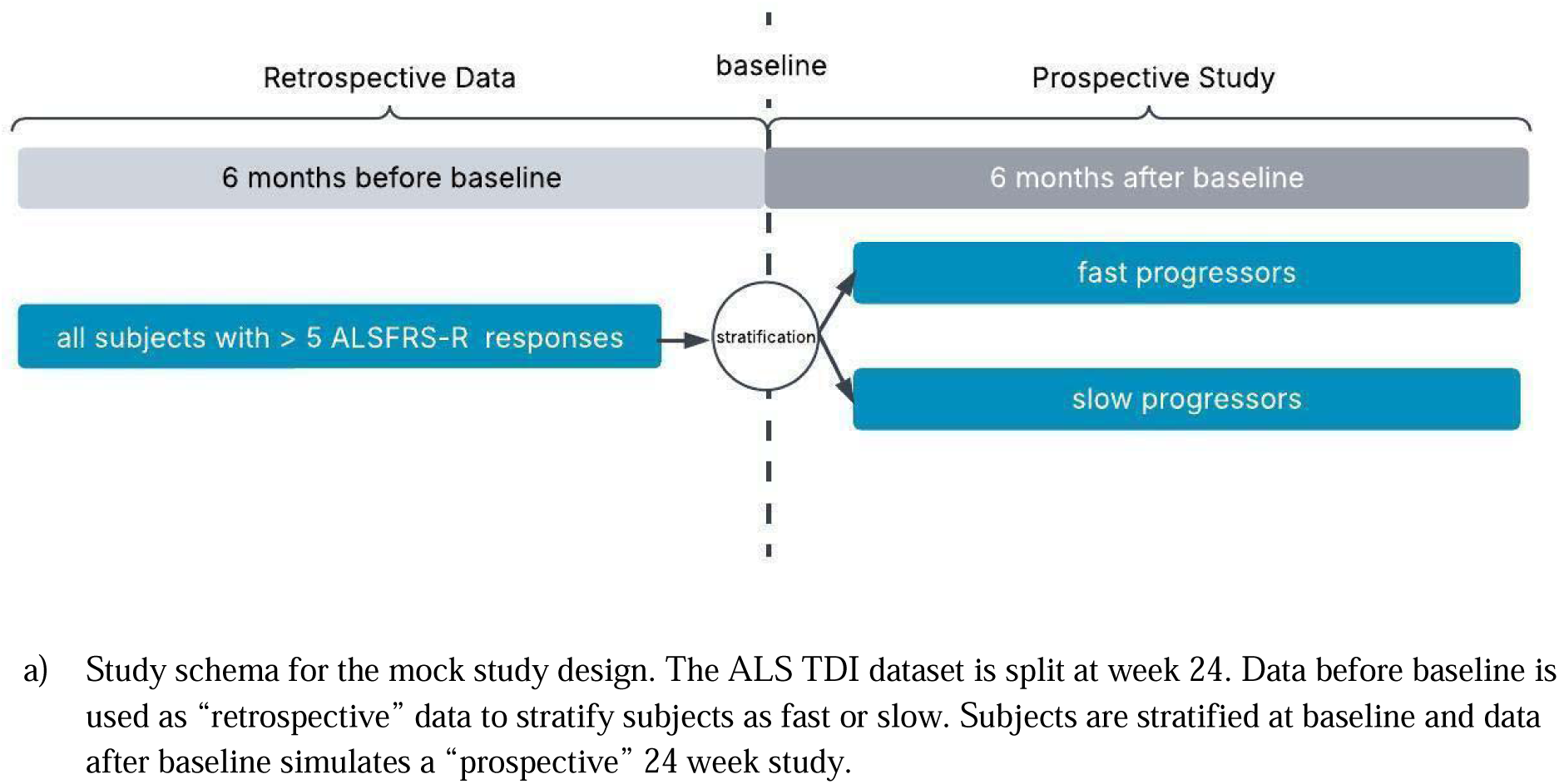
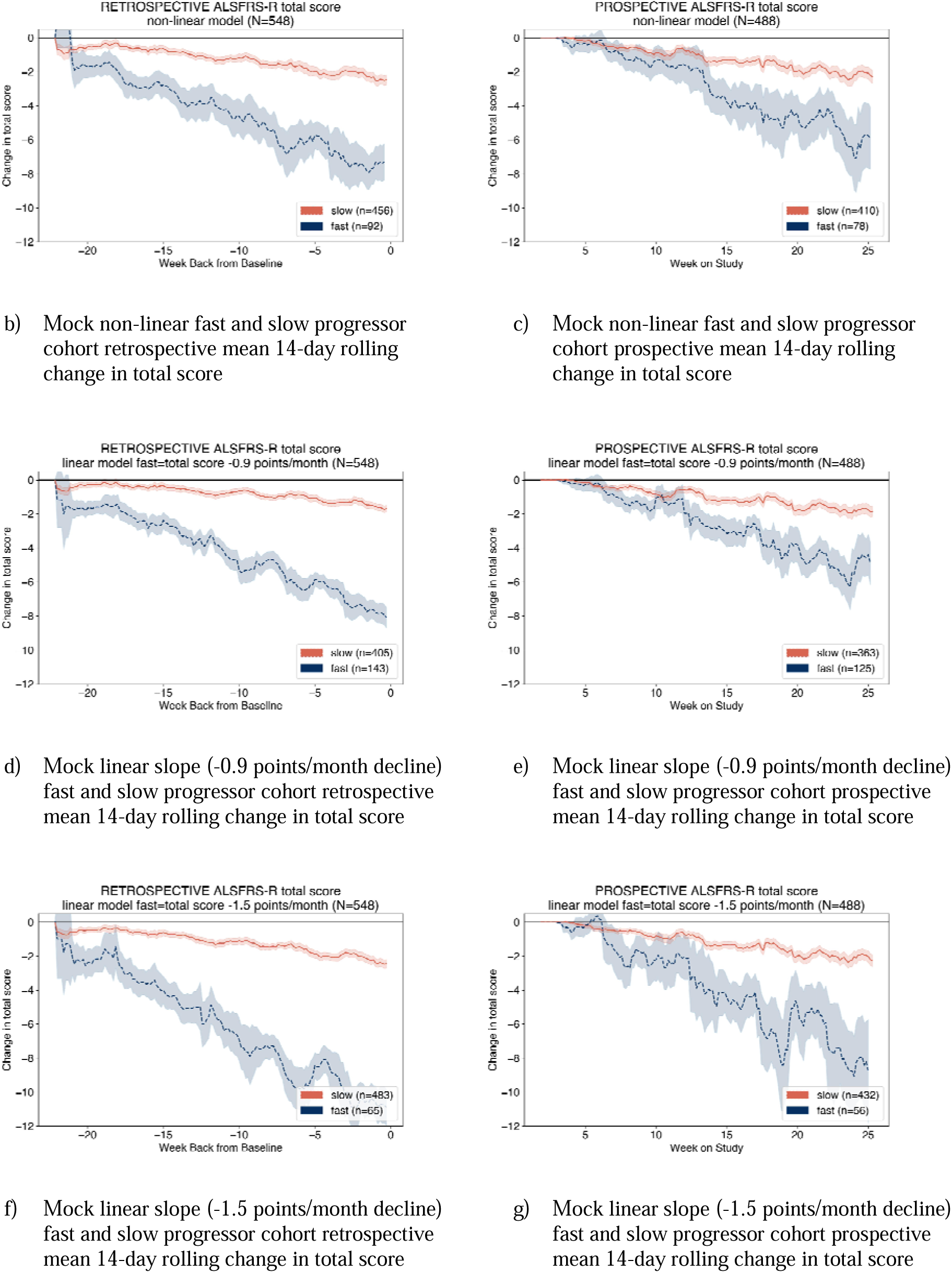
ALS TDI dataset simulation of a mock study with subjects stratified into fast and slow progressor cohorts using data from week zero to week 24 then prospective from weeks 24 (baseline) to 48 (end of study). The non-linear method is applied to ALSFRS-R total score change from data from week zero to 24 re-baselined as week minus 24 to week 0. Figures b and c show the retrospective and prospective change in total score with the non-linear model, figures d and e show the linear model with fast progressor slope threshold of −0.9 points/month, and figures f and g show the linear model with fast progression slope threshold of −1.5 points/month.

Using the non-linear model, fast progressor mean change [CI] in ALSFRS-R total score was −7.5 [−13.3,−2.4] points in the “retrospective” dataset and −6.8 [−12.0,−2.6] points in the “prospective” dataset. Slow progressor mean change in ALSFRS-R total score was −2.5 [−7.0,0.5] points in the “retrospective” dataset and −2.2 [−9.2,2.0] points in the “prospective” dataset.

When comparing linear and non-linear models for stratification, different proportions of subjects were classified as fast and slow (Supplemental Table 3). In a linear model with a fast progressor slope threshold of −1.5 points/month decline in ALSFRS-R total score, 65 (12%) were classified as fast, and a slope threshold of −0.9 points/month classified 143 (26%) as fast. The non-linear model classified 92 (17%) subjects as fast.

Fast progressors defined by the non-linear model showed more stable and consistent progression rates when comparing the mean change in ALSFRS-R total scores in the “retrospective” versus “prospective” datasets, with a mean change of 0.8 points/month between the 2 datasets. (Figures 5b to 5c, Supplemental Table 4). On the other hand, linear models estimated larger differences in decline in ALSFRS-R total score between the “retrospective” and “prospective” periods, with a total score mean difference of 4.2 points (Supplemental Table 4b) and 1.4 points (Supplemental Table 4c) than the non-linear model (Supplemental Table 4a). Similar patterns were observed for subdomain score changes and between fast and slow progressor cohorts (Figure 5b and c versus 5d and 5g, Supplemental Table 4).

## Discussion

Longitudinal ALSFRS-R scores over periods of at least 24 weeks in two datasets showed that mean percent change was greater in fine and gross motor subdomain scores than in total score and bulbar and respiratory subdomain scores, and that scores exhibited a non-linear trajectory of functional decline of ALS patients. In accounting for varying change patterns (fast, slow, alternating, or no progression), the nonlinear model characterized ALSFRS-R score progression better than a linear model, preserving a lower mean difference between mock retrospective and prospective cohorts. These results indicate that, as measured by the ALSFRS-R, ALS progresses nonlinearly to varying degrees across four functional domains.

Our findings are largely consistent with previous studies reporting that the ALSFRS-R measures three or four individual subdomains of disease [8, 9,10, 28, 29], and not purely a total score of function/disability for people living with ALS. At least two of these studies were also conducted with the PRO-ACT database [10,29] and others applied Rasch analysis [12], an approach that a priori requires a measure to exhibit unidimensionality. The observation that subdomains progress at a non-uniform rate has been indicated elsewhere [29,11] and was confirmed by our results. Past studies concluded that although ALSFRS-R effectively captures disease progression, there are opportunities to improve its measurement properties [10,11].

Differences in subdomain score probability distributions highlight the multi-dimensional, subdomain-specific nature of ALS disease progression. Additionally, the distributions suggest that respiratory and bulbar function may be preserved longer than fine and gross motor functions in studies up to 24 weeks in length and earlier in disease progression. Tofersen open label extension results reported by Biogen similarly showed that disease progression was detected more robustly in fine and gross motor than in bulbar and respiratory subdomains [30]. The use of change from baseline in ALSFRS-R total score as a clinical trial endpoint may mask actual treatment effects at the subdomain level. Therefore, these results reinforce the importance of subdomain-specific analysis in detecting ALS progression and sensitivity to treatment.

Because total score, and respiratory and bulbar subdomain scores, were skewed towards less severe symptoms and impacts at baseline, both datasets may represent participants who were relatively early in disease progression, potentially reflecting a sub-population of ALS patients targeted by interventional therapies or able to participate in a clinical study due to less advanced disease. This finding highlights the need to develop more sensitive measures of ALS progression than the ALSFRS-R earlier in disease, especially for bulbar and respiratory functions. At present, studies examining progression in or treatment for early ALS using the ALSFRS-R may benefit from focusing on changes in fine and gross motor function.

The nonlinearity of disease progression has implications for clinical practice and for the expectations of patients and their caregivers. The model presented in this work preserves observed features of disease progression on an individual basis and does not average them over the duration of observation. These findings reinforce the heterogenous nature of ALS, including the possibility that some symptoms and functions may improve for some patients over the short term despite long term deterioration. Non-linearity is a critical issue for the stratification of participants in clinical trials. Linear progression models estimating prerandomization ALSFRS-R slope may have led in part to inconclusive results [31]. Stratification has several benefits, including the potential to conduct primary analysis in a subset of study participants that is defined based on clinical characteristics such as rapid progression, with secondary or supportive analyses of the full study sample [6]. Because stratification is based on the anticipated progression patterns of specific participants, such patterns must be characterized with optimal accuracy. The analysis of a mock study in this work demonstrated that a nonlinear model is superior to a linear model and should be applied to stratification of trial participants when ALSFRS-R scores are used. In a clinical setting, a four progression cohort definition may be unnecessary detail and alternative fast/slow cohort definition used. In a two progression cohort design, fast subjects are defined by the fast progressor definition in this work and slower subjects as all other subjects. A two cohort definition has the advantage of classifying progression in all subjects because the classifier is binary and was implemented in the mock study (Figure 5).

Our study benefits from several methodological strengths that enhance confidence in the findings. The nonlinear model was developed on an aggregate longitudinal interventional dataset of ALS subjects taking placebo and then tested on registry data of subjects who volunteered for a natural history study. This largely avoids study specific limitations of smaller sample size models. This approach demonstrates that the model applies in both supervised clinical trial and real-world settings. Moreover, the non-linear model presented uses simple thresholds rather than machine learning to identify fast progressors, which may support greater interpretability of the results.

Several limitations should be kept in mind when interpreting these findings. Both of the datasets used in analysis are based on uneven data collection schedules that resulted in a nonuniform cadence and quantity of ALSFRS-R score samples over time. In addition, the nonlinear model we developed was tested on aggregate data rather than a single study, though these analyses could be performed on a single study dataset given a sufficient number of ALSFRS-R scores collected before and after baseline. Finally, the non-linear model requires a sufficient number of observations per subject: sensitivity testing suggests that a minimum of five ALSFRS-R visit samples is required to conclusively classify the progression type of more than 75% of subjects in a dataset.

Future research should seek to further improve the measurement properties of the ALSFRS-R, and to identify additional tools and methods to measure ALS disease progression. A general clinical measure of ALS such as the Rasch Overall ALS Disability Scale (ROADS) holds promise as a unidimensional patient-reported outcome measure developed using modern item response theory [11]. Patients provided input on the initial ROADS item bank, but no published literature to date indicates whether the final ROADS was cognitively debriefed with patients. Therefore, more research is needed to further establish the content validity of ROADS along with studies to quantify its sensitivity to meaningful change in severity of disability.

Alternatively, domain-specific clinical outcome measures (based on patient relevance and hypothesized treatment benefit), biomarkers, and measures using sensor-based digital health technology (sDHT) also merit further exploration. Seeking to comprehensively characterize disease severity and progression more accurately than a total score, the ALS Impairment Multidomain Scale (AIMS) was developed as a Rasch-built PRO with unidimensional subscales for bulbar, motor and respiratory [32]. Neurofilament light chain (NfL) is a marker of neuronal injury that is significantly elevated in ALS. Because higher NfL levels are associated with more rapid disease progression and predict shorter survival, this biomarker may be incorporated in stratification approaches to help identify faster progressing patients. sDHT offers the potential to measure disease-relevant motor function (e.g. physical activity, gait, upper extremity function, speech) in a manner that could be more sensitive, frequent, and representative of real-world function than traditional measures. Based on our findings that motor subdomains show greater longitudinal change than bulbar and respiratory subdomains, sDHT-based assessments of fine and gross motor function may yield changes that are more readily observed [33,34]. If used instead to supplement the less sensitive subdomains of the ALSFRS-R, sDHT-based assessments of participants’ speech [35] swallowing, or breathing could detect subtle changes that may not be observed otherwise.

Given the mounting evidence of ALS disease heterogeneity and progression variability, a reconceptualization of the ALSFRS-R may prove useful. A non-linear approach to disease progression, a prioritization of assessing fine and gross motor subdomains in early stages of disease, and a novel definition for fast and slow progressors may optimize study designs and lead to more efficient endpoints in ALS. Future work exploring the relationship between alternative measures and ALSFRS-R, as well as the best way to integrate and combine these measures, may contribute to more sensitive and efficient ALS clinical trials and speed the development of new therapies.

## Data Availability

All data produced in the present study are available upon reasonable request to the authors

https://www.als.net/arc/data-commons/

https://ncri1.partners.org/PROACT

## Contributions

Design: R.E., A.S. / Analysis R.E., A.S., M.W., D.T., J.L., S.M., W.N. / Manuscript R.E., W.N., O.L., M.W., J.W.

## Acknowledgements

The authors wish to thank Brittany Gentile, Regeneron, for her input regarding confirmatory factor analysis.

## References

1. Brown, R.H. & Al-Chalabi, A. Amyotrophic lateral sclerosis. N. Engl.J. Med. 377, 162–172 (2017).

2. Hardiman, O., et al. (2017). “Amyotrophic lateral sclerosis.” Nat Rev Dis Primers 3: 17071.

3. Cedarbaum, J. M., et al. (1999). “The ALSFRS-R: a revised ALS functional rating scale that incorporates assessments of respiratory function. BDNF ALS Study Group (Phase III).” J Neurol Sci 169(1-2): 13–21

4. Rofail, D., et al. (2025). “Advancing Future Amyotrophic Lateral Sclerosis Medicines by Incorporating The Patient Voice Into Patient-Centered Holistic Measurement Strategies for Clinical and Real-World Studies: Results from Targeted Literature Reviews.” (submitted)

5. EMA (2015). “Guideline on clinical investigation of medicinal products for the treatment of amyotrophic lateral sclerosis (ALS)” Document Number EMA/531686/2015, Corr.1. (link) last accessed March 2025

6. FDA (2019). “Amyotrophic Lateral Sclerosis: Developing Drugs for Treatment Guidance for Industry - Guidance for Industry” Docket Number FDA-2013-N-0035. (link) last accessed January 2025

7. Miano, B., et al. (2004). “Inter-evaluator reliability of the ALS functional rating scale.” Amyotroph Lateral Scler Other Motor Neuron Disord 5(4): 235–239.

8. Bakker, L. A., et al. (2017). “Assessment of the factorial validity and reliability of the ALSFRS-R: a revision of its measurement model.” J Neurol 264(7): 1413–1420.

9. van Eijk, R. P. A., et al. (2021). “An old friend who has overstayed their welcome: the ALSFRS-R total score as primary endpoint for ALS clinical trials.” Amyotroph Lateral Scler Frontotemporal Degener 22(3-4): 300–307.

10. Young, C. A., et al. (2024). “Improving the measurement properties of the Amyotrophic Lateral Sclerosis Functional Rating Scale-Revised (ALSFRS-R): deriving a valid measurement total for the calculation of change.” Amyotroph Lateral Scler Frontotemporal Degener 25(3-4): 400–409.

11. Fournier, C. N., et al. (2020). “Development and Validation of the Rasch-Built Overall Amyotrophic Lateral Sclerosis Disability Scale (ROADS).” JAMA Neurol 77(4): 480–488.

12. Bakers, J. N. E., et al. (2022). “Using the ALSFRS-R in multicentre clinical trials for amyotrophic lateral sclerosis: potential limitations in current standard operating procedures.” Amyotroph Lateral Scler Frontotemporal Degener 23(7-8): 500–507.

13. Din Abdul Jabbar, M. A., et al. (2024). “Predicting amyotrophic lateral sclerosis (ALS) progression with machine learning.” Amyotroph Lateral Scler Frontotemporal Degener 25(3-4): 242–255.

14. Kueffner, R., et al. (2019). “Stratification of amyotrophic lateral sclerosis patients: a crowdsourcing approach.” Sci Rep 9(1): 690.

15. Witzel, S., et al. (2022). “Fast versus slow disease progression in amyotrophic lateral sclerosis-clinical and genetic factors at the edges of the survival spectrum.” Neurobiol Aging 119: 117–126.

16. Gordon, P. H., et al. (2010). “Progression in ALS is not linear but is curvilinear.” Journal of Neurology 257(10): 1713–1717. Frontotemporal Degener 24(1-2): 108–116.

17. Ramamoorthy, D., et al. (2022). “Identifying patterns in amyotrophic lateral sclerosis progression from sparse longitudinal data.” Nat Comput Sci 2(9): 605–616.

18. Hamatani, T., et al. (2024). “ALSFRS-R decline rate prior to baseline is not useful for stratifying subsequent progression of functional decline.” Amyotroph Lateral Scler Frontotemporal Degener 25(3-4): 388–399.

19. Din Abdul Jabbar, M. A., et al. (2024). “Describing and characterizing variability in ALS disease progression.” Amyotroph Lateral Scler Frontotemporal Degener 25(1-2): 34–45.

20. Atassi, N., et al. (2014). “The PRO-ACT database: design, initial analyses, and predictive features.” Neurology 83(19): 1719–1725.

21. Bakker, L. A., et al. (2020). “Development and assessment of the inter-rater and intra-rater reproducibility of a self-administration version of the ALSFRS-R.” Journal of Neurology, Neurosurgery & Psychiatry 91(1): 75–81.

22. Benatar, M., et al. (2024). “Rethinking phase 2 trials in amyotrophic lateral sclerosis.” Brain.

23. FDA (2022). RADICAVA ORS label claim. Reference ID: 4982904 (revised May 2022). (link) last accessed March 2025

24. Cedarbaum, J. M., et al. (1999). “The ALSFRS-R: a revised ALS functional rating scale that incorporates assessments of respiratory function. BDNF ALS Study Group (Phase III).” J Neurol Sci 169(1-2): 13–21.

25. Castrillo-Viguera, C., et al. (2010). “Clinical significance in the change of decline in ALSFRS-R.” Amyotroph Lateral Scler 11(1-2): 178–180.

26. Verma, A. (2021). Clinical Manifestation and Management of Amyotrophic Lateral Sclerosis. Amyotrophic Lateral Sclerosis. T. Araki. Brisbane (AU), Exon Publications

27. Kong, Q., et al. (2020). “Python Programming and Numerical Methods - A Guide for Engineers and Scientists.” 1st Edition ISBN 9780128195499

28. Franchignoni, F., et al. (2013). “Evidence of multidimensionality in the ALSFRS-R Scale: a critical appraisal on its measurement properties using Rasch analysis.” Journal of Neurology, Neurosurgery & Psychiatry 84(12): 1340–1345.

29. Bacci, E. D., et al. (2016). “Item response theory analysis of the Amyotrophic Lateral Sclerosis Functional Rating Scale-Revised in the Pooled Resource Open-Access ALS Clinical Trials Database.” Amyotrophic Lateral Sclerosis and Frontotemporal Degeneration 17(3-4): 157–167.

30. Biogen MA Inc., QALSODY® (tofersen) Briefing document. NDA# 215887 Peripheral and Central Nervous System Drugs Advisory Committee. Meeting date 22 March 2023.

31. Miller, T. M., et al. (2022). “Trial of Antisense Oligonucleotide Tofersen for SOD1 ALS.” N Engl J Med 387(12): 1099–1110.

32. de Jongh, A. D., et al. (2023). “Development of a Rasch-Built Amyotrophic Lateral Sclerosis Impairment Multidomain Scale to Measure Disease Progression in ALS.” Neurology 101(6): e602–e612.

33. Acien, A., et al. (2024). “A novel digital tool for detection and monitoring of amyotrophic lateral sclerosis motor impairment and progression via keystroke dynamics.” Sci Rep 14(1): 16851

34. Gupta, A. S., et al. (2023). “At-home wearables and machine learning sensitively capture disease progression in amyotrophic lateral sclerosis.” Nat Commun 14(1): 5080.

35. Stegmann, G., et al. (2024). “Automated speech analytics in ALS: higher sensitivity of digital articulatory precision over the ALSFRS-R.” Amyotrophic Lateral Sclerosis and Frontotemporal Degeneration 25(7-8): 767–775.

