## Supplemental Tables and Figures for "Driving novel endpoints and study designs in amyotrophic lateral sclerosis: closer examination of the ALSFRS-R subdomains and a new definition of fast and slow progressors"

Supplemental tables and figures are listed in the order in which they are referenced.

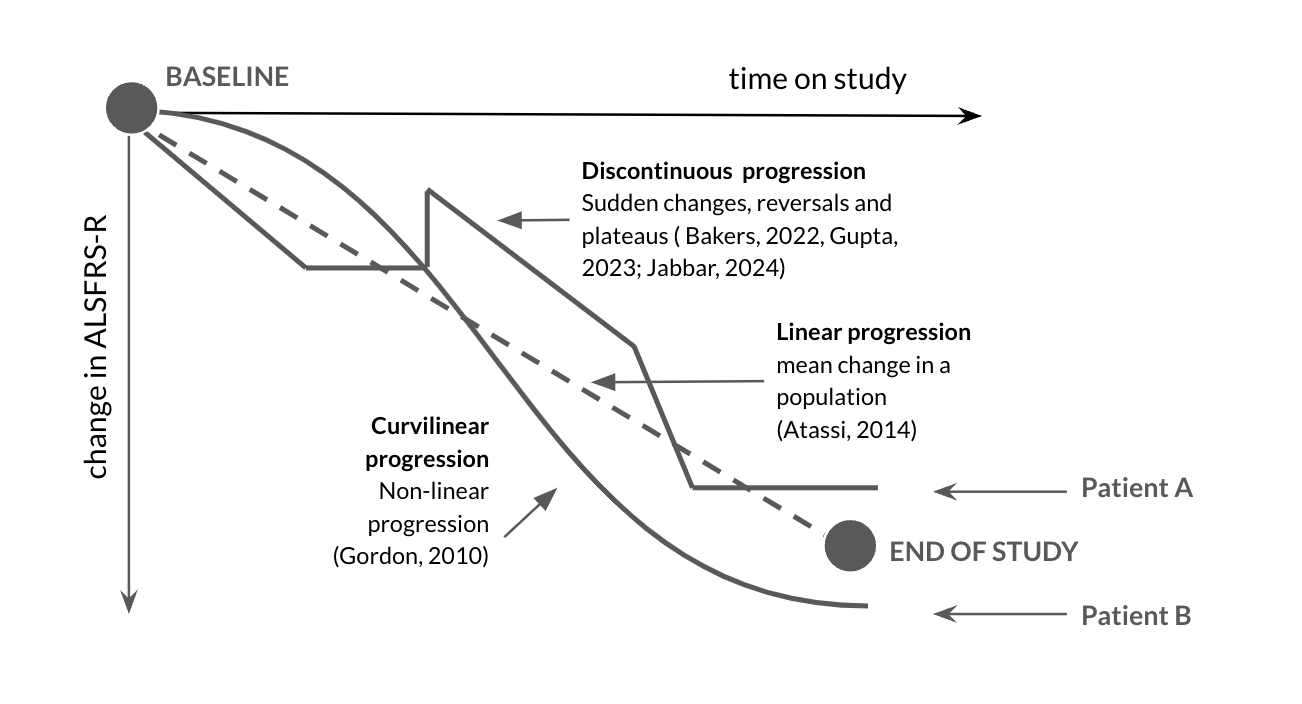

**Supplemental Figure 1**. **Illustrative progression pathways for different observed patient progression pathways versus linear progression.** Linear progression between baseline and end of study is plotted with a dashed line. Solid lines represent different non-linear progression profiles observed in ALSFRS-R data with related citations. Linear progression overestimates progression for Patient A and underestimates for Patient B. Citations indicate reported types of progression in the literature.

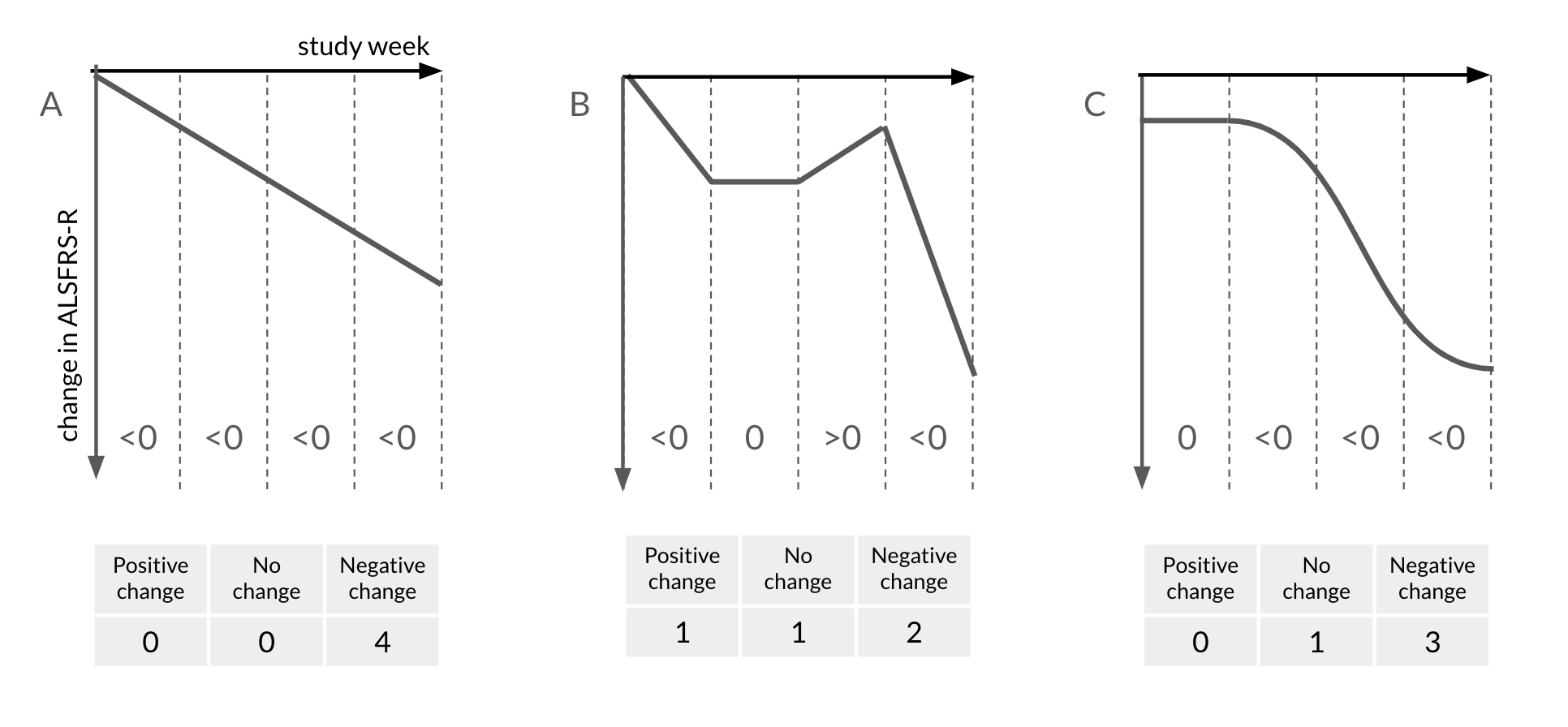

**Supplemental Figure 2. Illustration of the finite difference method adapted for use with ALS progression.**  Three types of progression profile from Figure 2 are illustrated: a) linear, b) alternating/reversals and c) curvilinear. By way of example, using subject B, change in ALSFRS-R is negative (<0) in the first period and flat (0) in the second period. The first period does not have a direction because there is no prior period. Inset tables show the count of each type of change by subject. Subjects B and C have approximately the same decline but different trajectories.

| 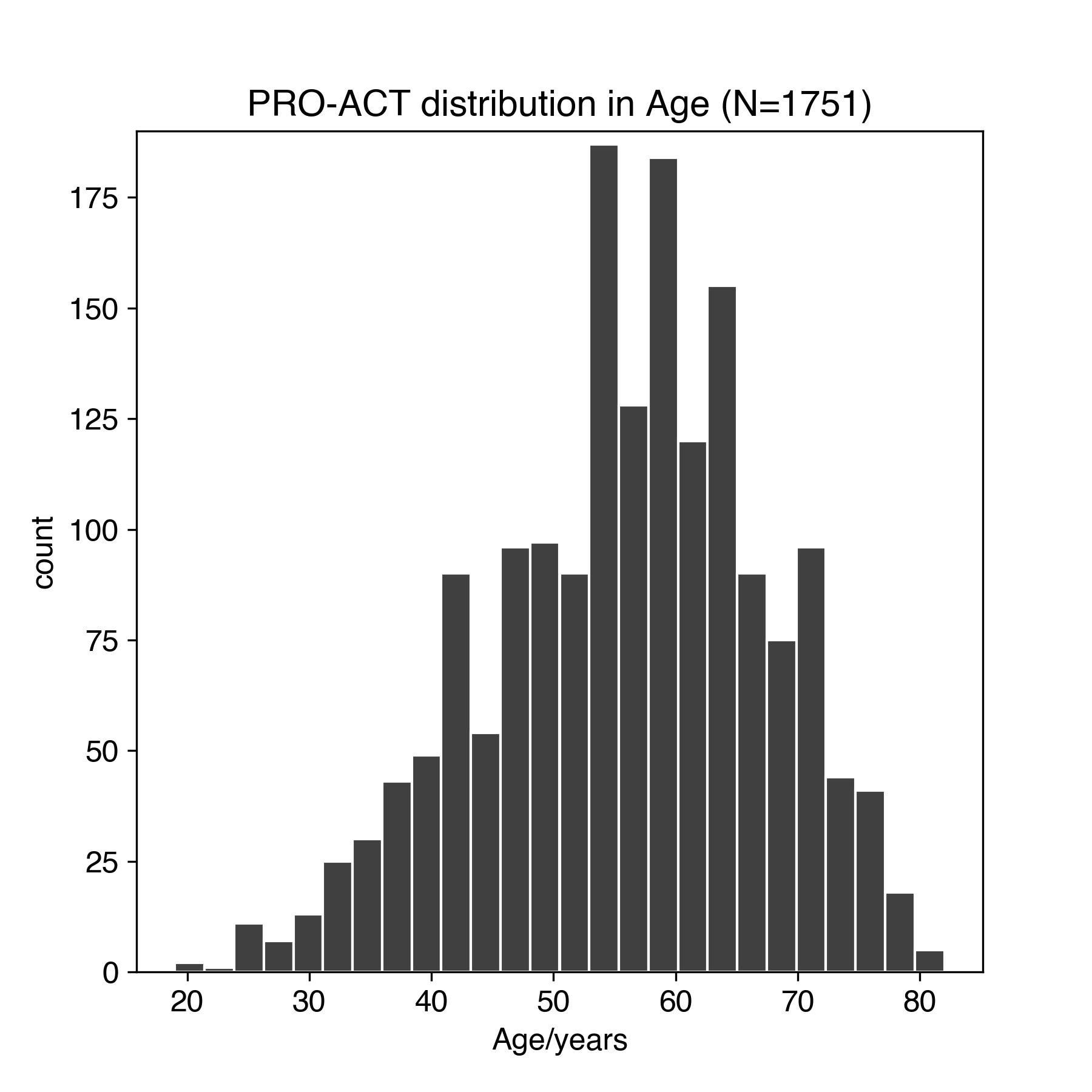 | 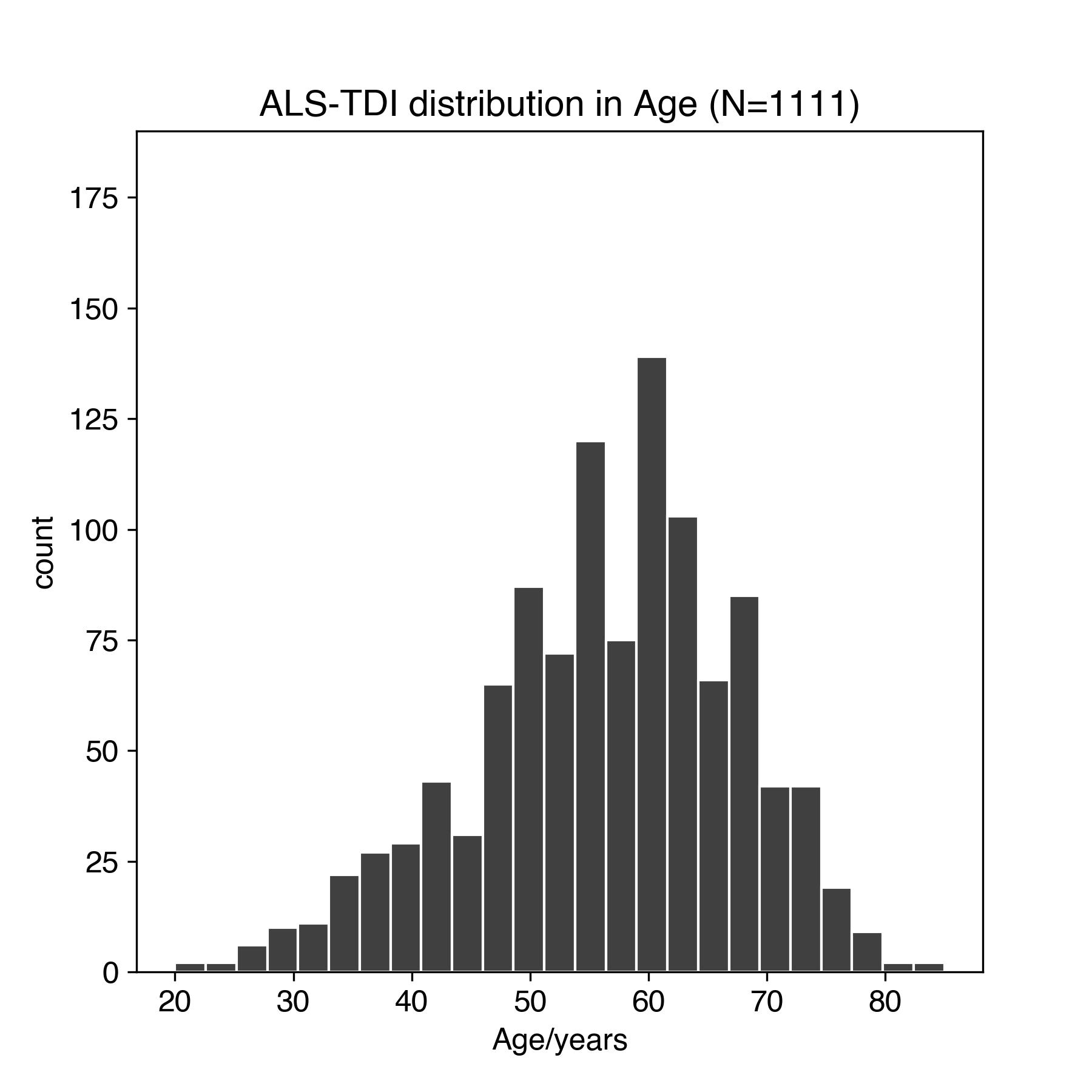 |
| --- | --- |
| a) PRO-ACT and ALS TDI subject age | |
| 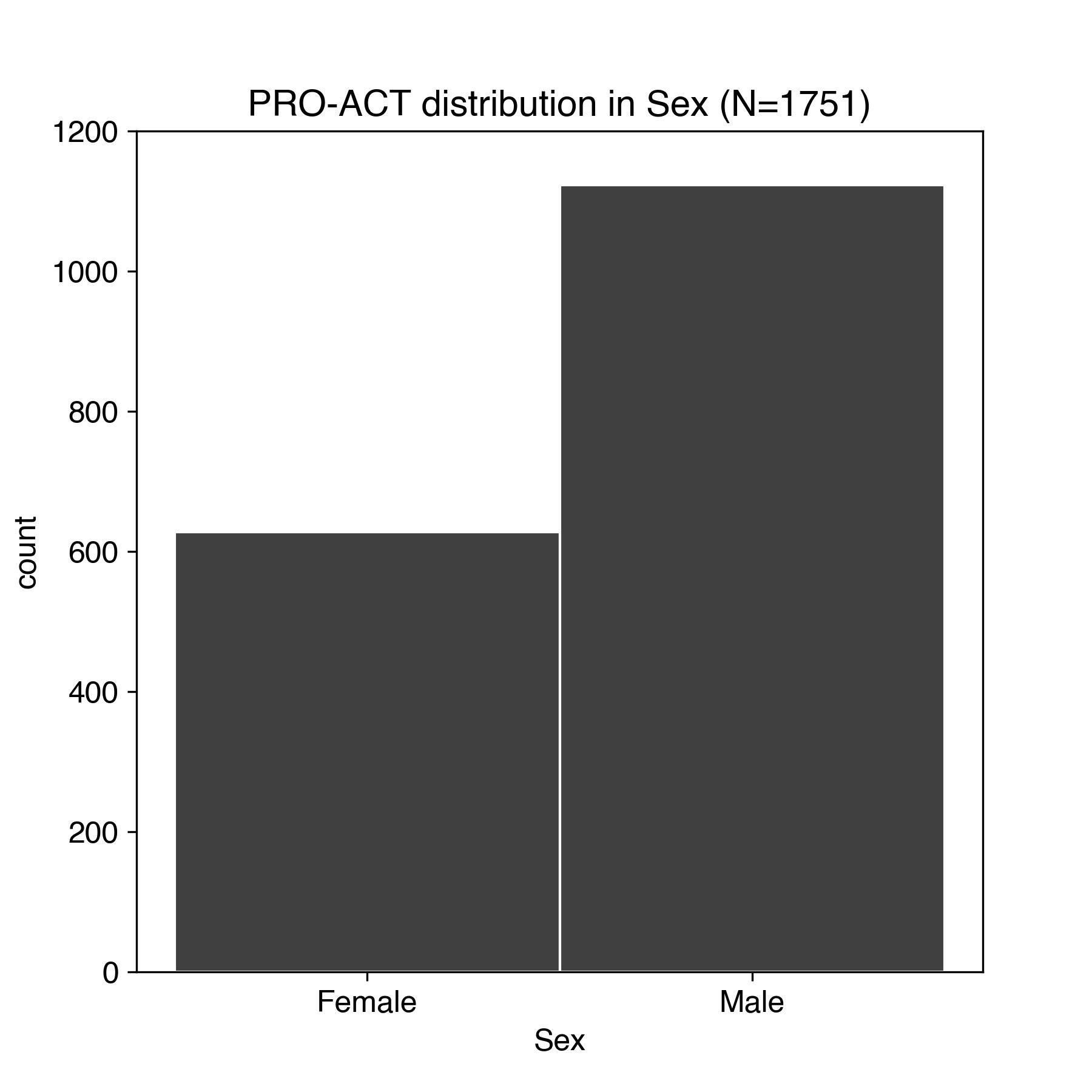 | 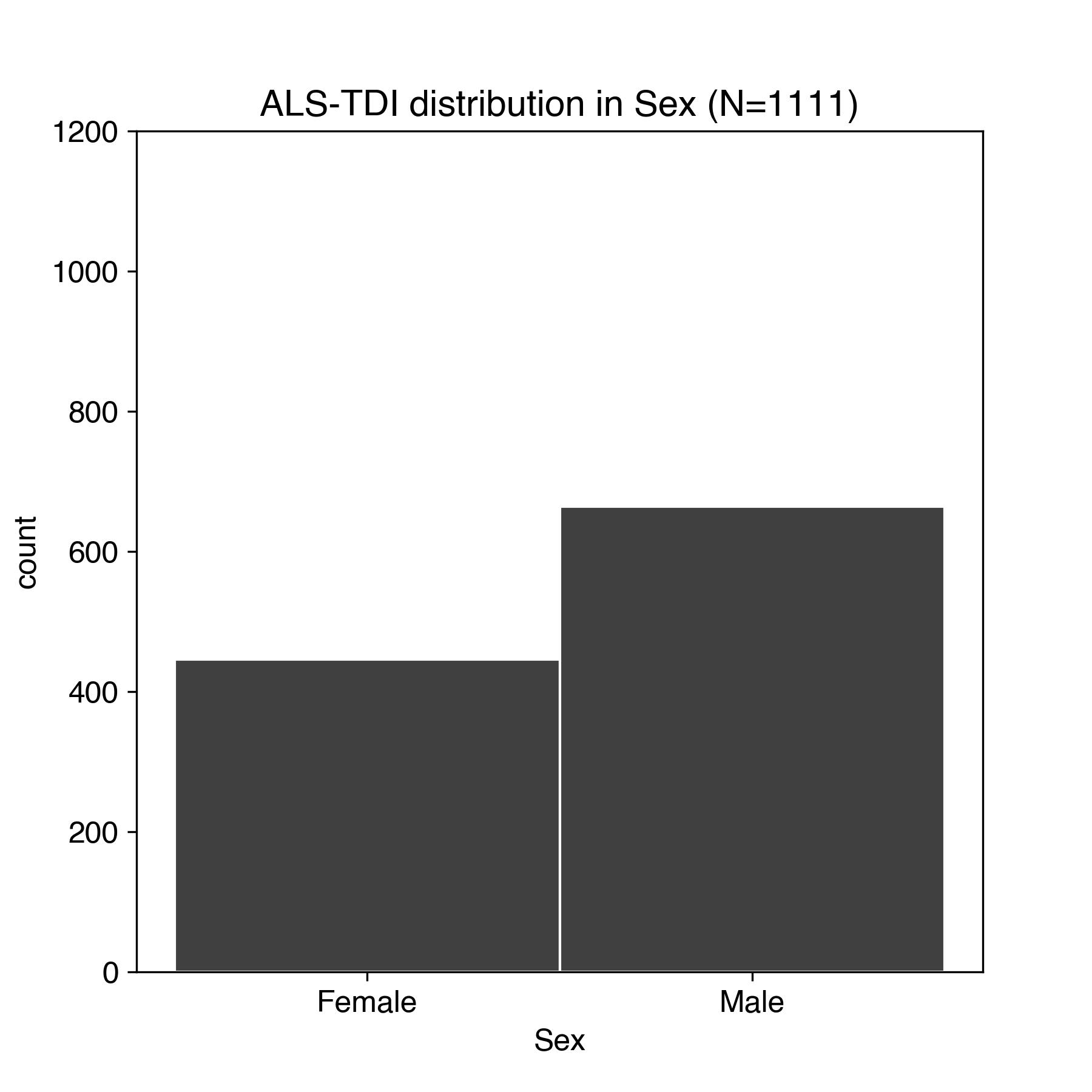 |
| b) PRO-ACT and ALS TDI subject sex | |
| 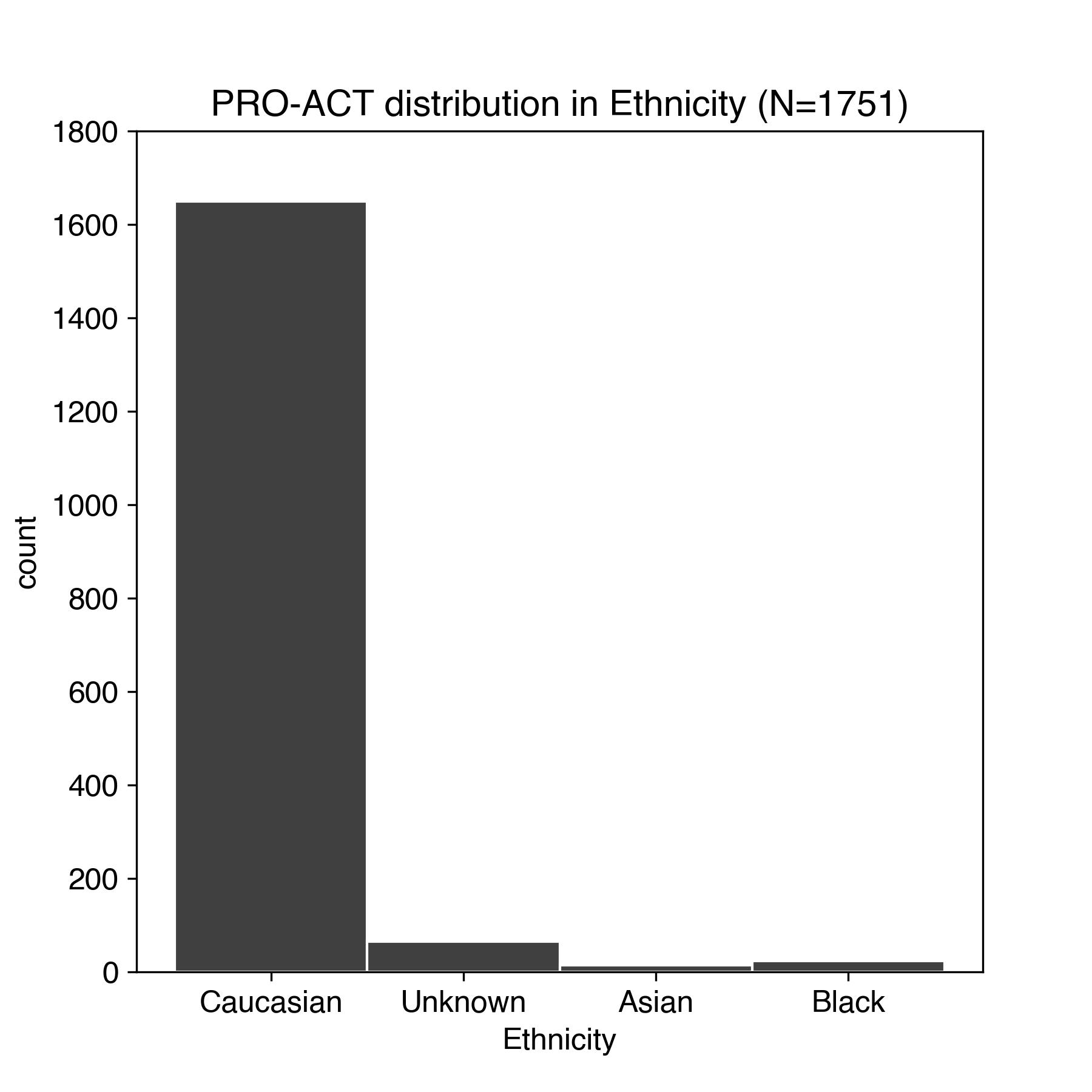 | 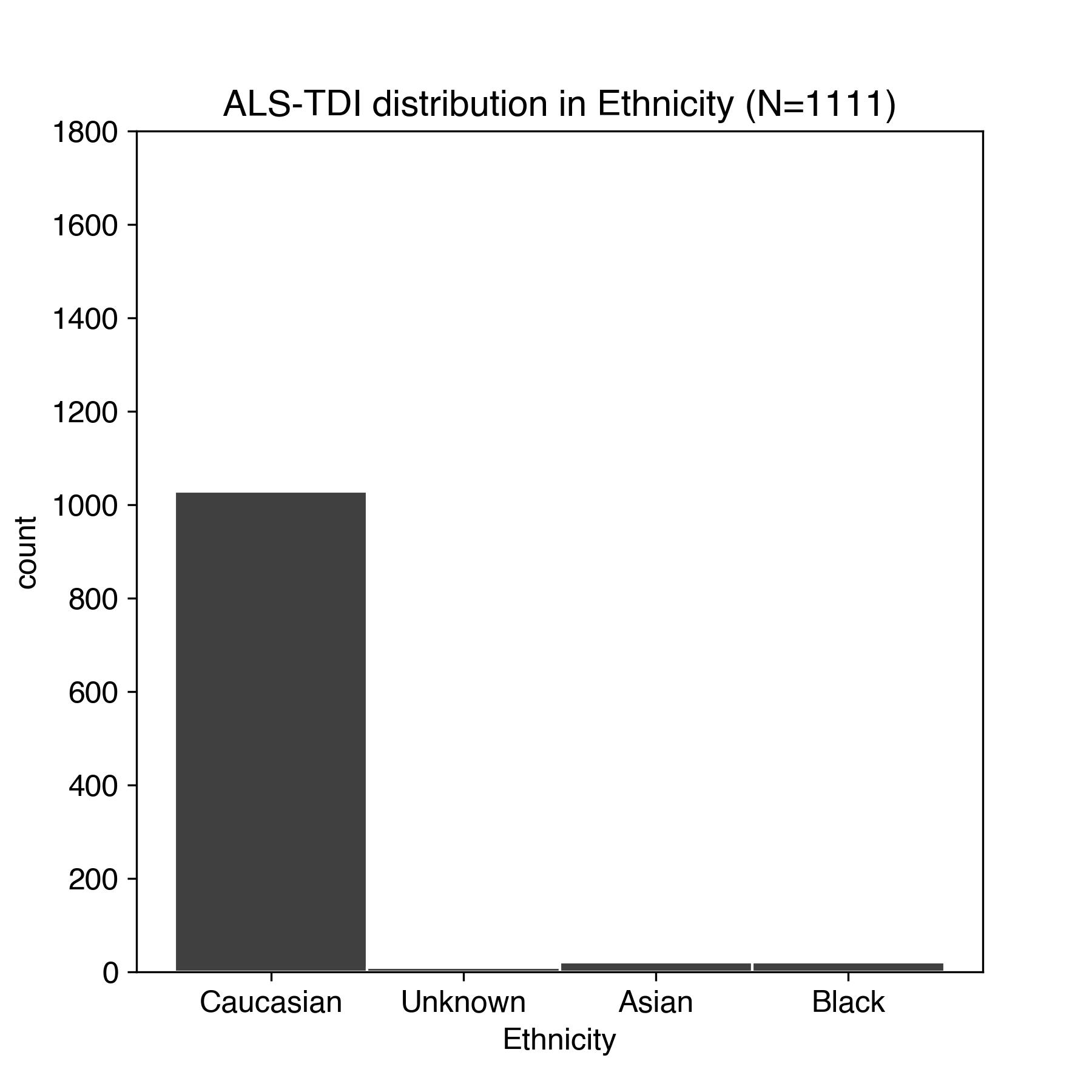 |
| c) PRO-ACT and ALS TDI subject ethnicity | |
| 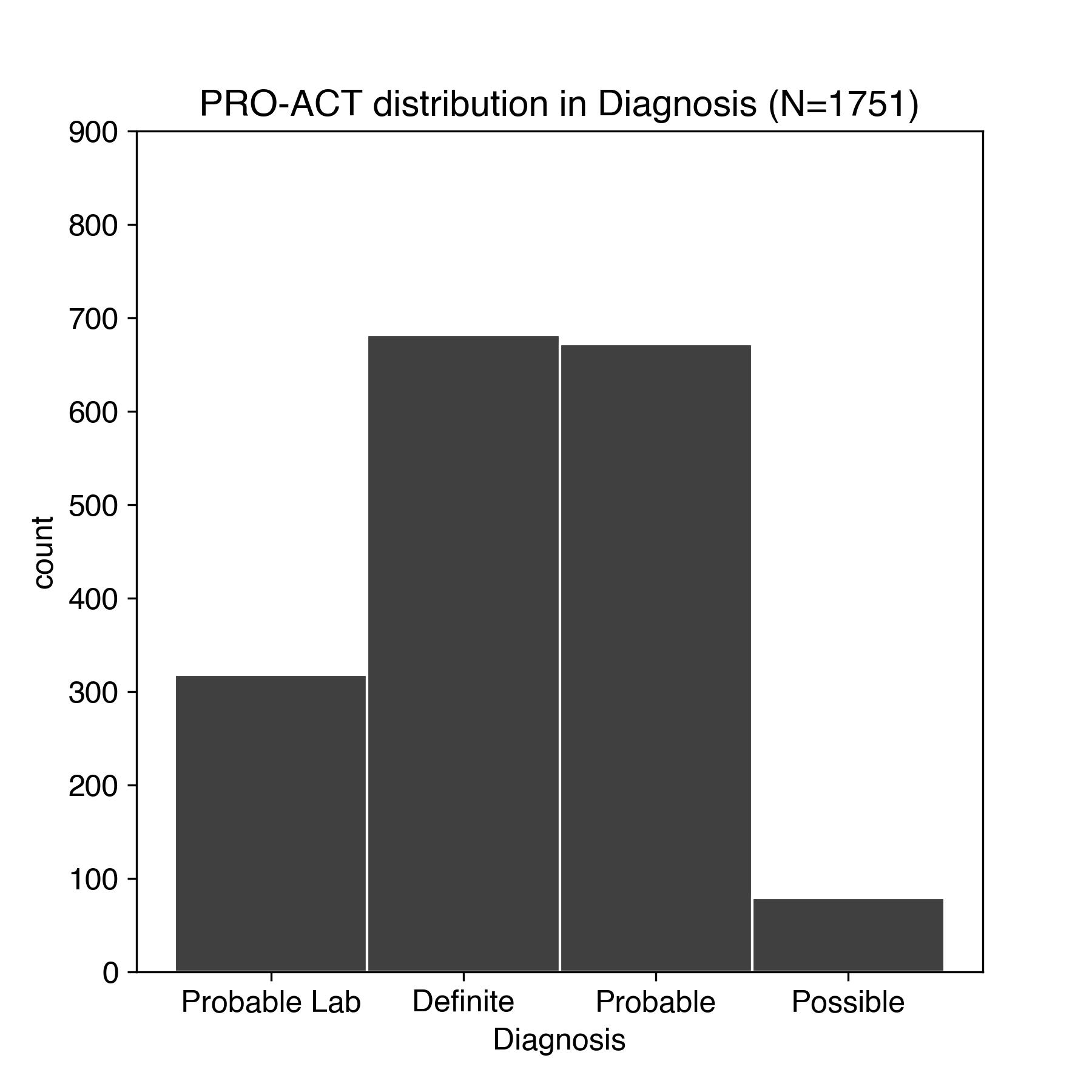 | 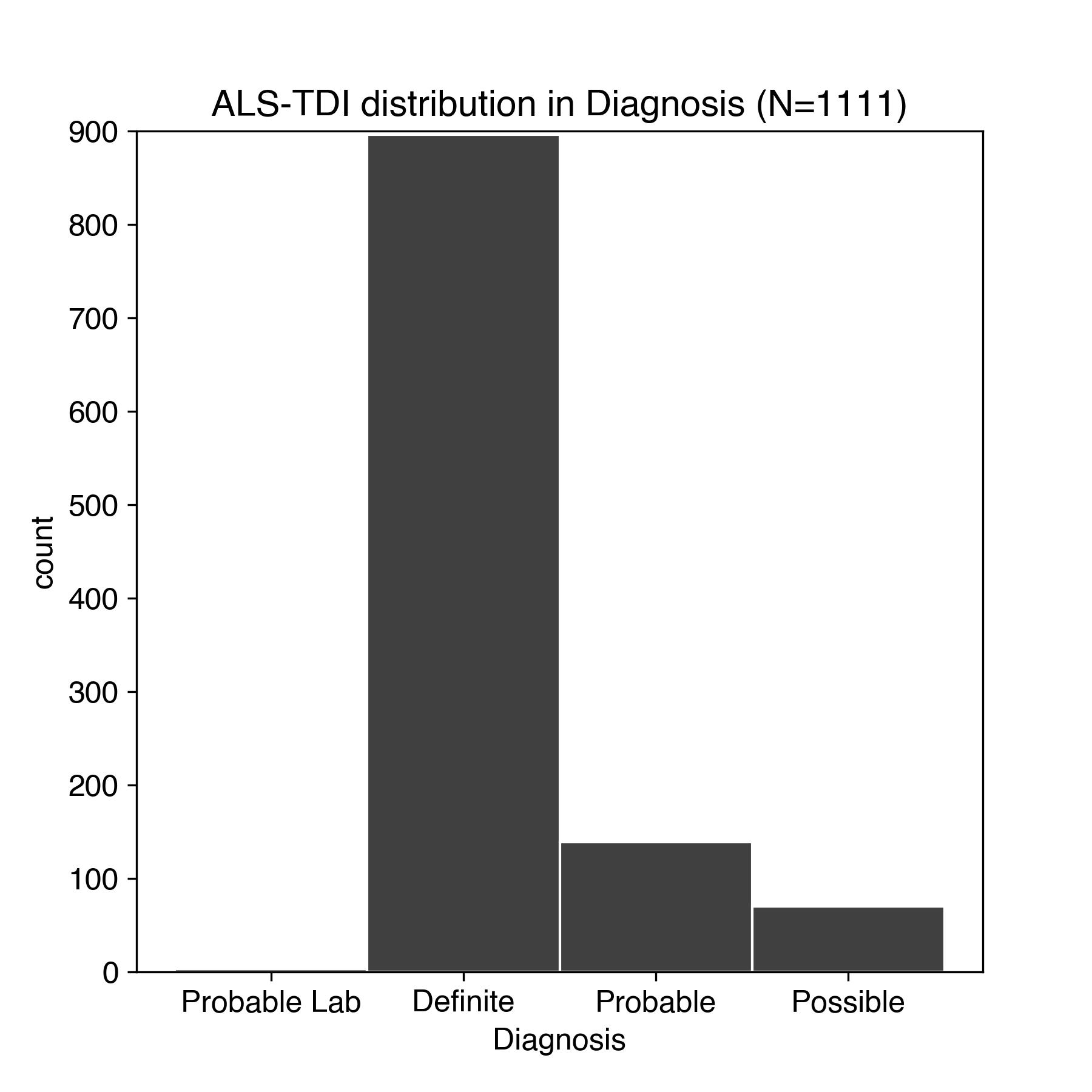 |
| d) PRO-ACT and ALS TDI distribution in onset location | |
| 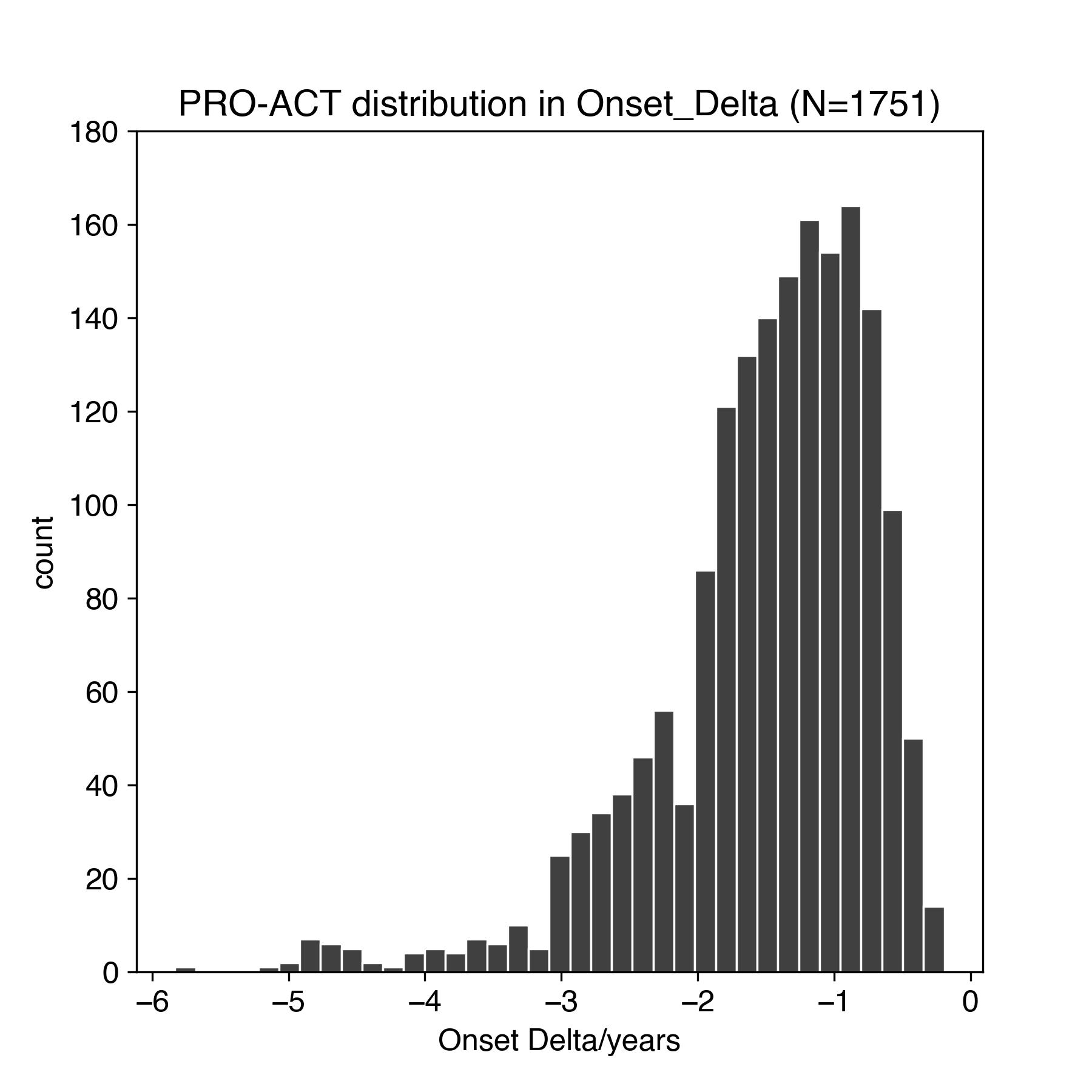 | 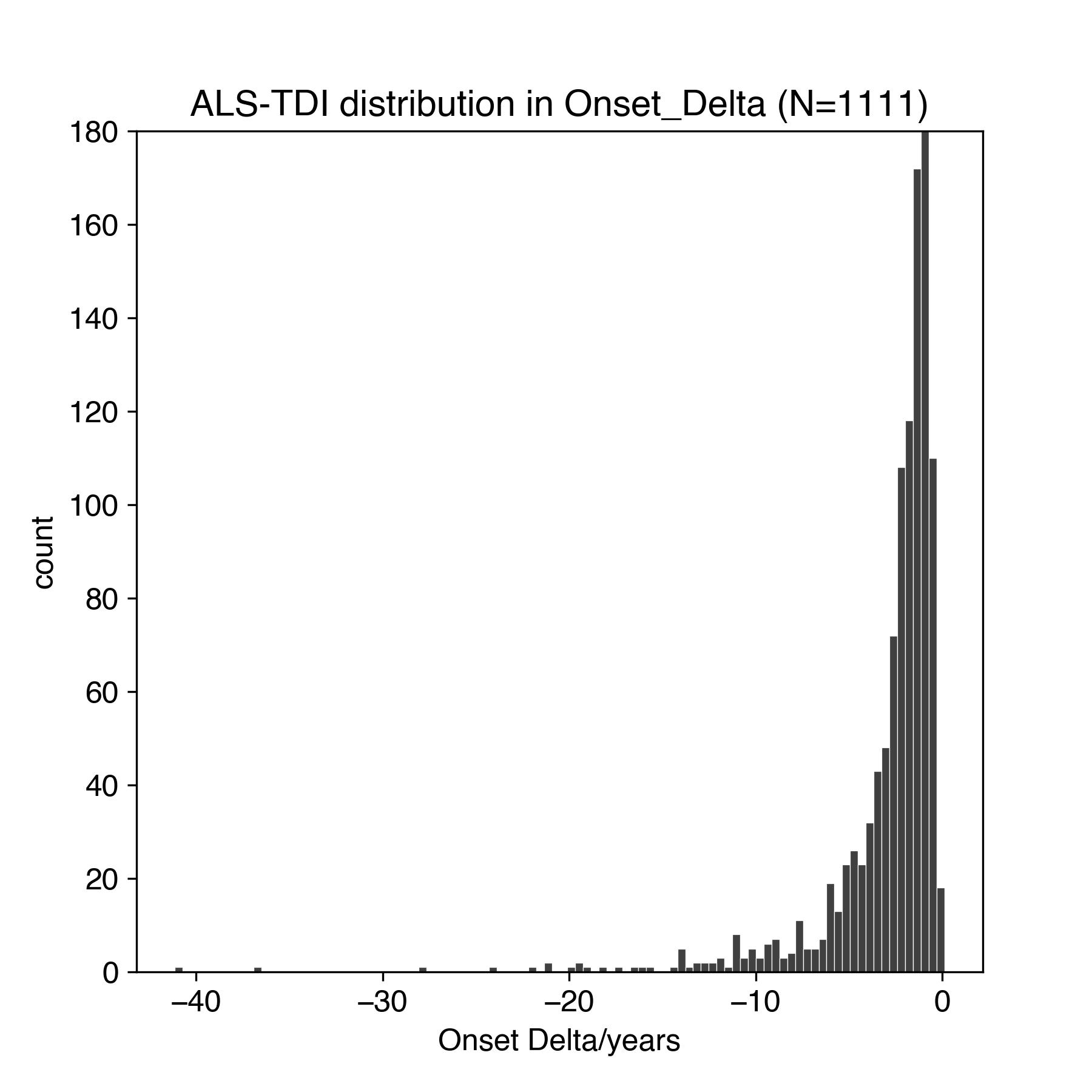 |
| e) PRO-ACT and ALS TDI subject onset delta - onset date of first symptoms in years back from study baseline. Note difference in x-axis scale between panels. | |
| 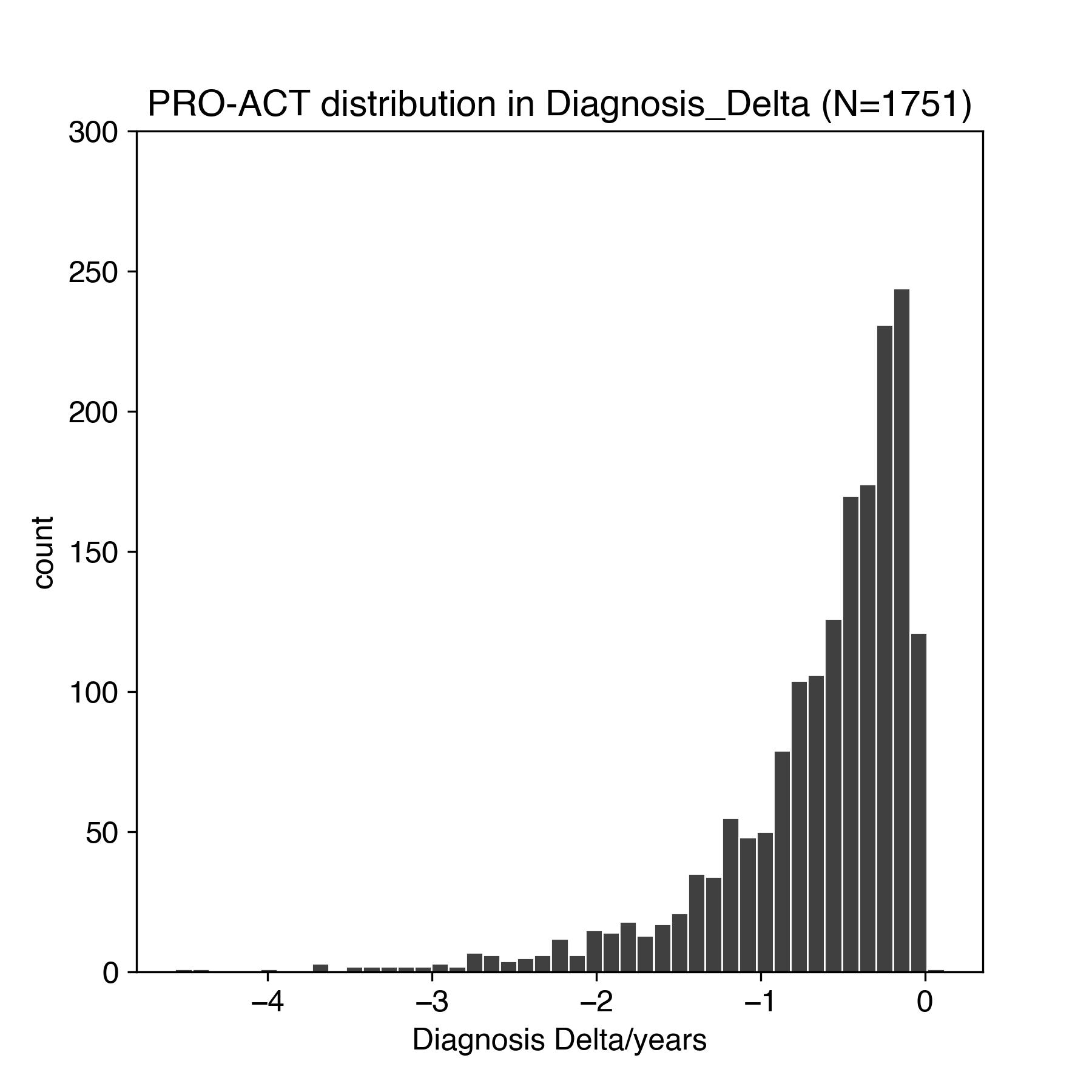 | 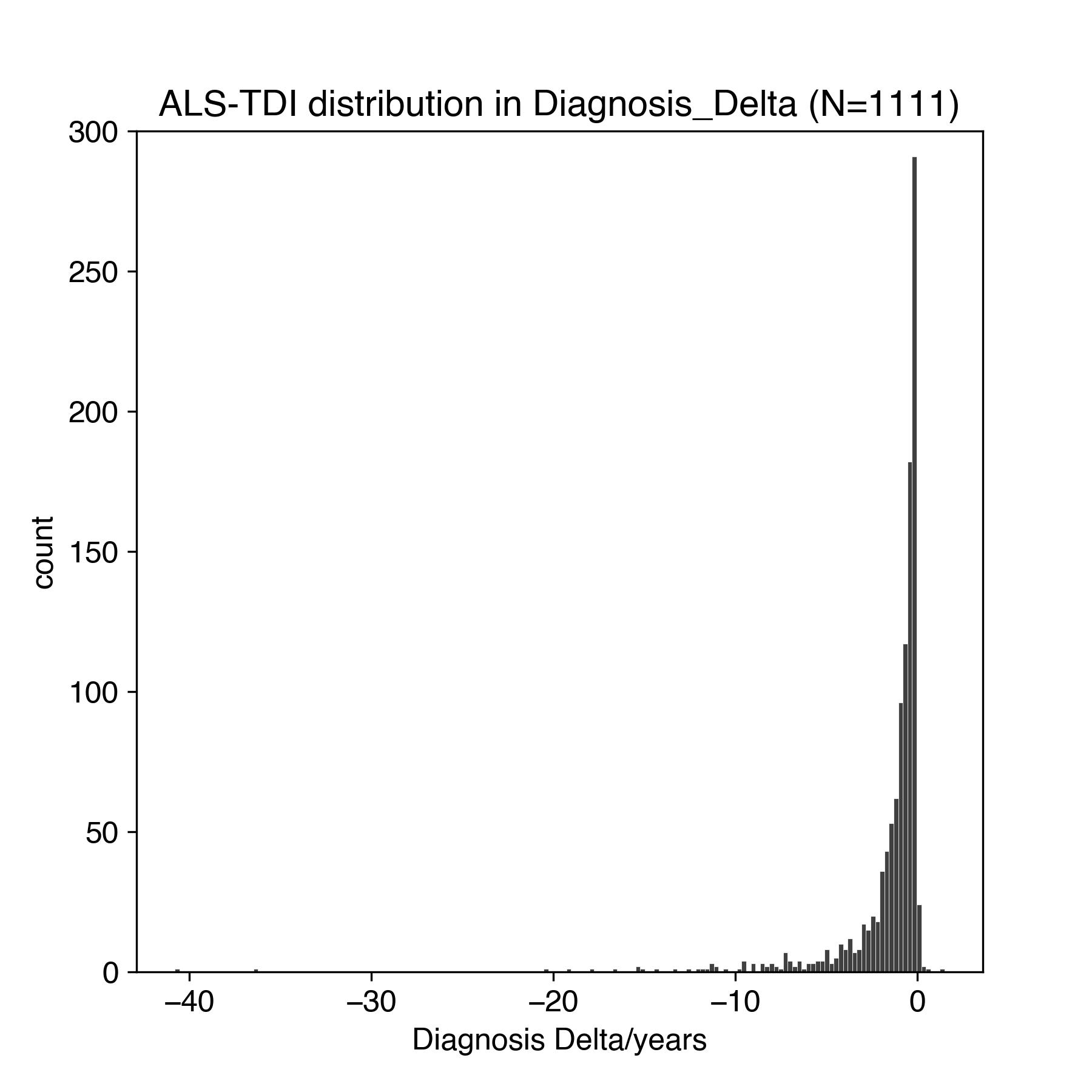 |
| f) PRO-ACT and ALS TDI subject diagnosis delta - diagnosis date as years back from baseline. Note difference in x-axis scale between panels. | |
| 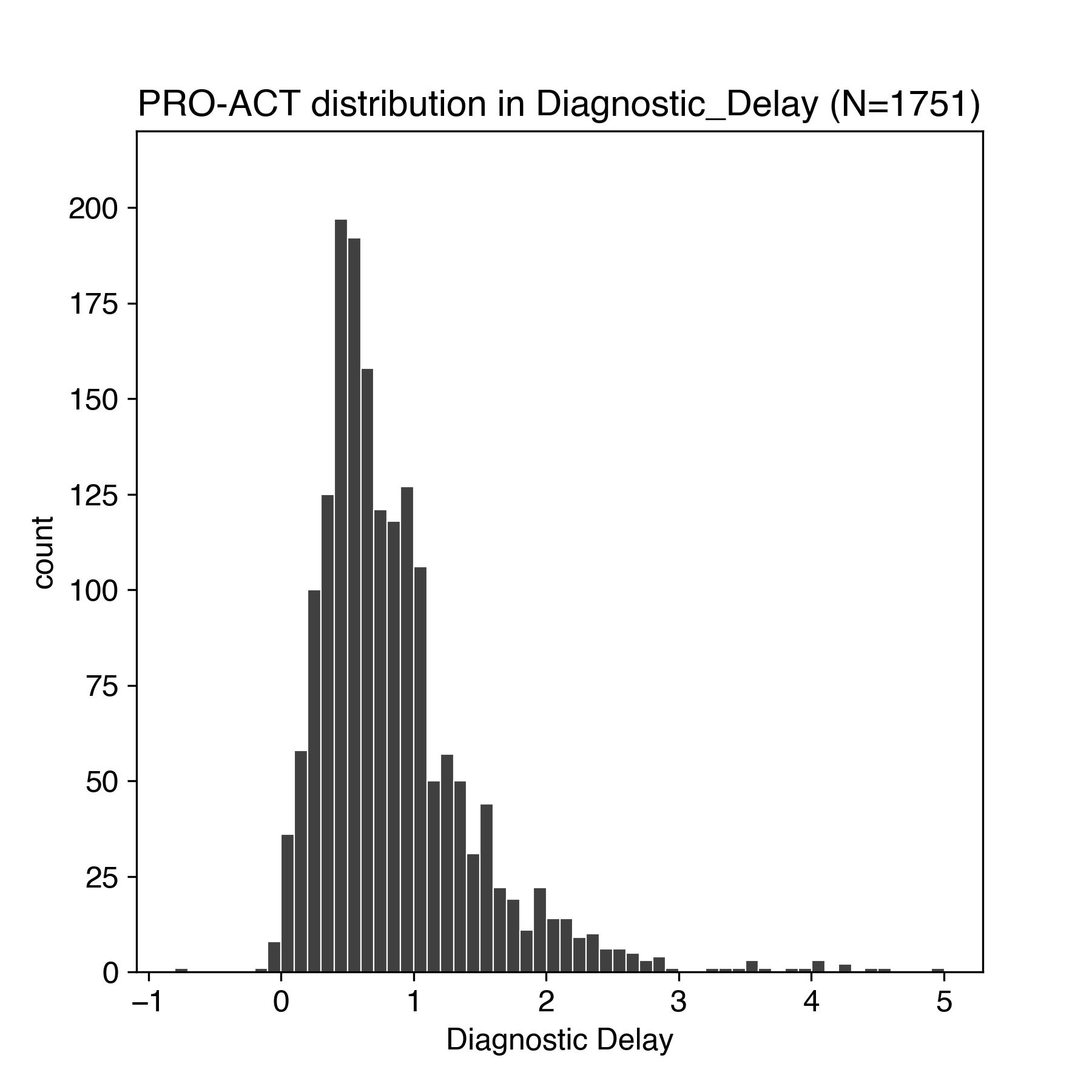 | 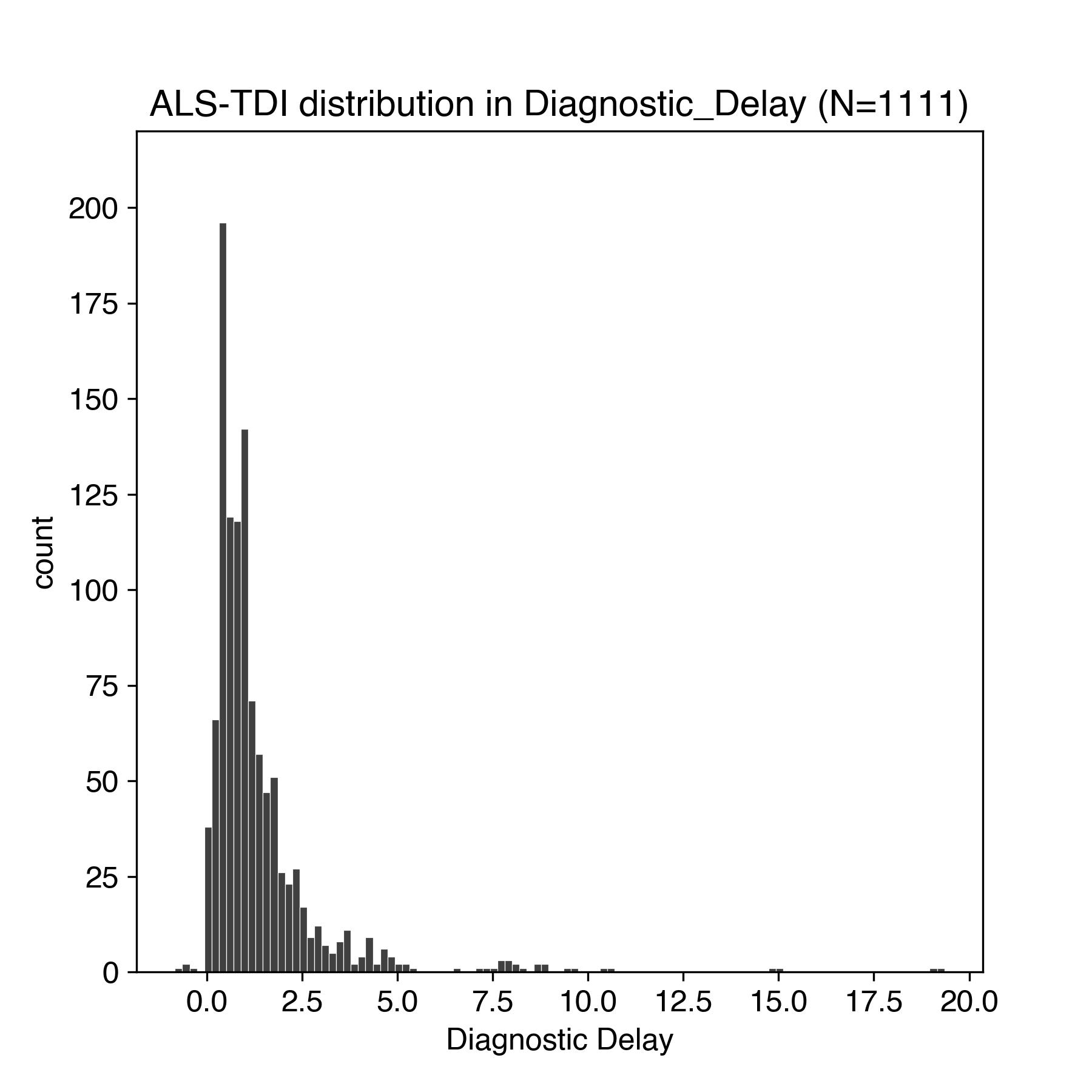 |
| g) PRO-ACT and ALS TDI subject diagnostic delta - difference in years between onset and diagnosis. Note difference in x-axis scale between panels. | |
| 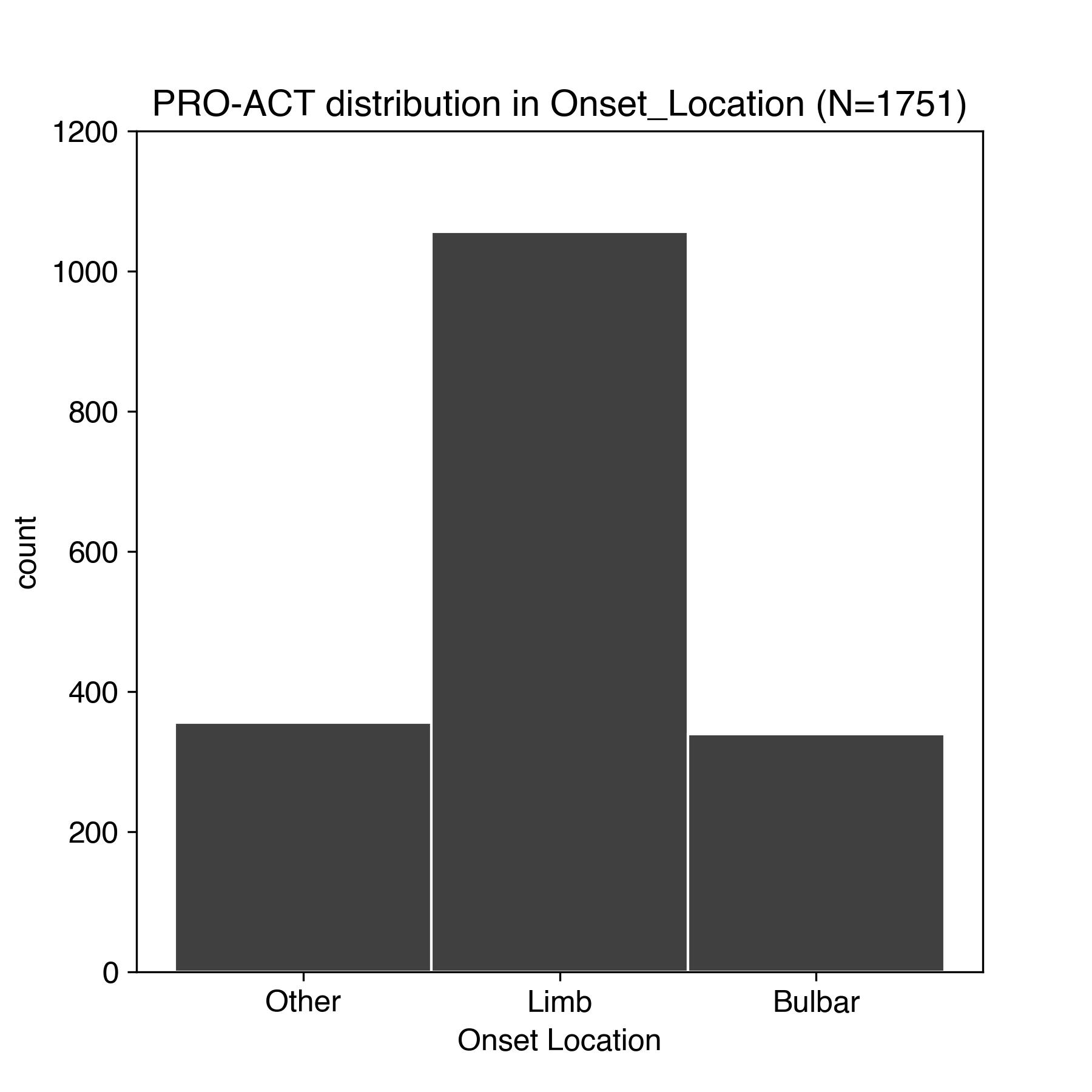 | 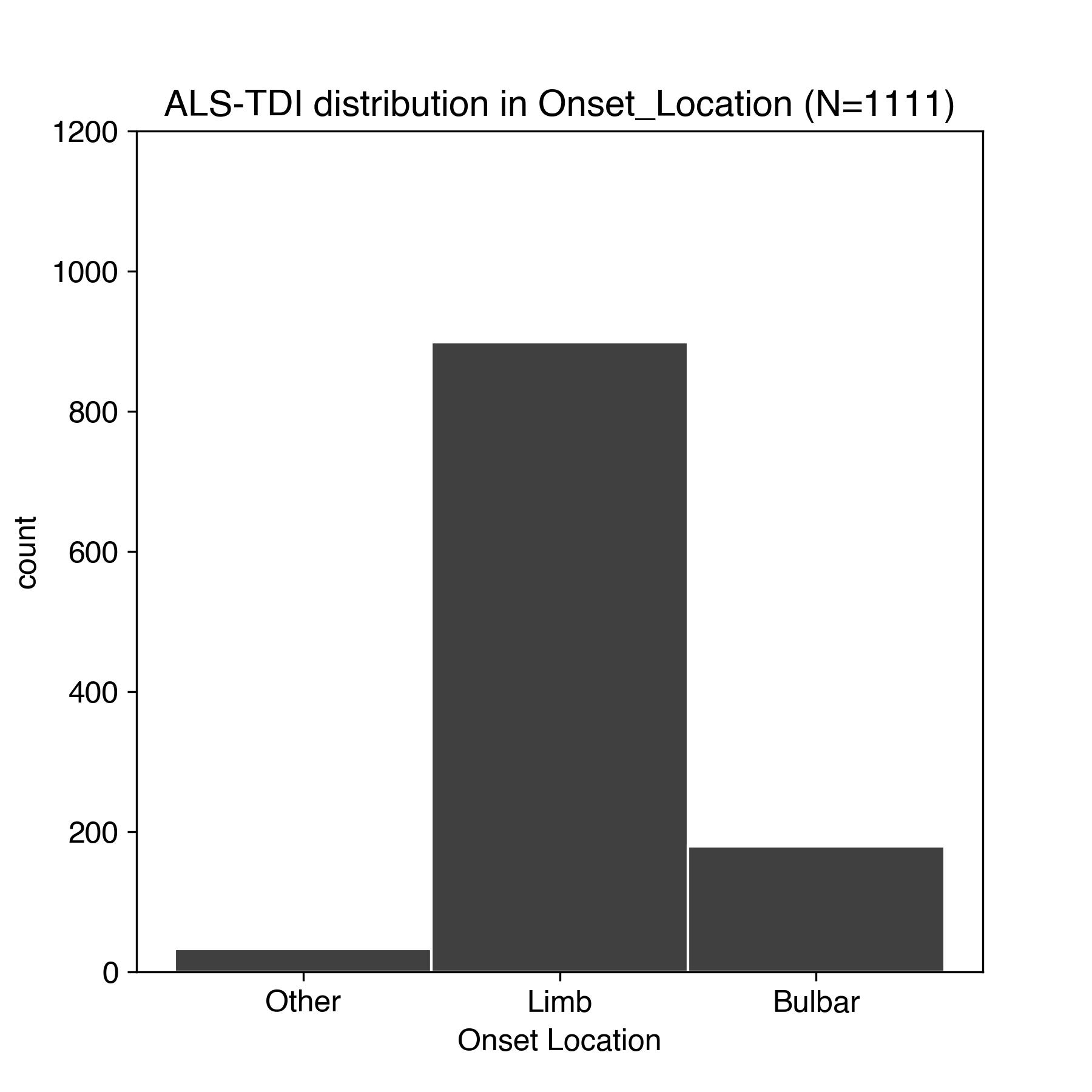 |
| h) PRO-ACT and ALS TDI subject disease onset location - the body area where symptoms were first observed | |

**Supplemental Figure 3. Histograms of each baseline measure in the PRO-ACT and ALS TDI dataset**. Variable names are plotted as labeled in the dataset and values plotted without curation such as definition of and removal of outliers. Diagnostic delay is calculated as diagnosis delta minus onset delta.

**Supplemental Table 1.** **Confirmatory factor structure.**  Indices are tabulated for each of the factor models examined: df: Degrees of Freedom; RMSEA: Root Mean Square Error of Approximation; TLI: Tucker-Lewis Index; CFI: Comparative Fit Index; AIC: Akaike Information Criterion; BIC: Bayesian Information Criterion

| **Factor Model** | **Chi-square** | **df** | **RMSEA** | **TLI** | **CFI** | **AIC** | **BIC** |
| --- | --- | --- | --- | --- | --- | --- | --- |
| 1-factor | 2572.9713 | 54 | 0.2774 | 0.0991 | 0.2629 | 39.5223 | 145.3270 |
| 3-factor | 903.6522 | 51 | 0.1661 | 0.6771 | 0.7505 | 51.0226 | 170.0528 |
| 4-factor | 499.9542 | 48 | 0.1246 | 0.8181 | 0.8677 | 58.3527 | 190.6086 |
| 3-factor + 1-factor | 113.9457 | 36 | 0.0598 | 0.9582 | 0.9773 | 83.6246 | 268.7828 |
| 4-factor + 1-factor | 85.3022 | 32 | 0.0524 | 0.9678 | 0.9844 | 91.7189 | 294.5113 |

a) PRO-ACT dataset placebo week zero confirmatory factor structure (N=792)

| **Factor Model** | **Chi-square** | **df** | **RMSEA** | **TLI** | **CFI** | **AIC** | **BIC** |
| --- | --- | --- | --- | --- | --- | --- | --- |
| 1-factor | 5316.2023 | 54 | 0.2628 | 0.4374 | 0.5397 | 40.4700 | 166.5363 |
| 3-factor | 2184.5890 | 51 | 0.1722 | 0.7585 | 0.8134 | 50.9057 | 192.7303 |
| 4-factor | 1306.8920 | 48 | 0.1363 | 0.8486 | 0.8899 | 58.1489 | 215.7318 |
| 3-factor + 1-factor | 887.8653 | 36 | 0.1295 | 0.8634 | 0.9255 | 82.7424 | 303.3584 |
| 4-factor + 1-factor | 139.5467 | 32 | 0.0488 | 0.9806 | 0.9906 | 91.8023 | 333.4294 |

b) ALS TDI dataset week zero confirmatory factor structure (N=1,111)

| 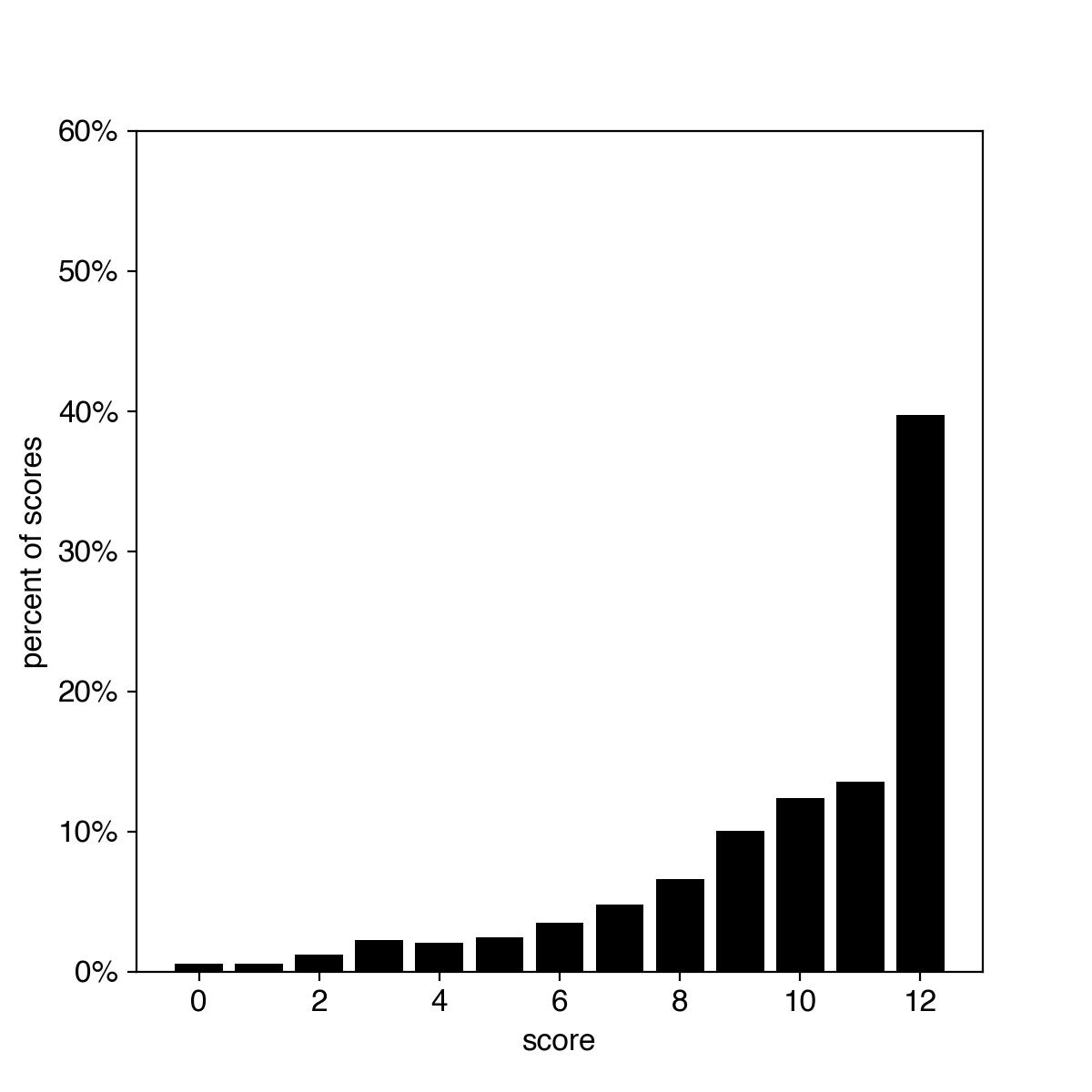 | 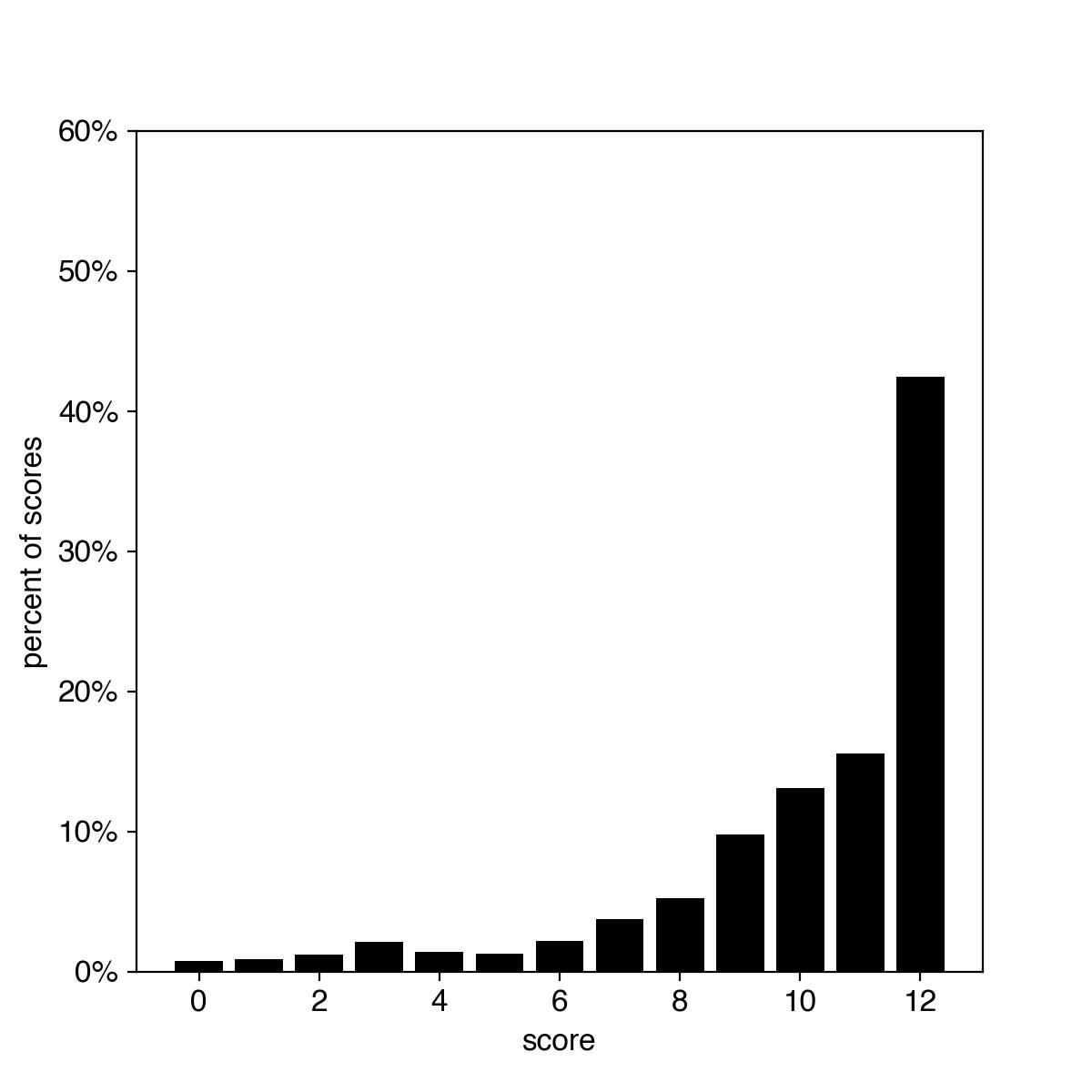 |
| --- | --- |
| a) Bulbar subdomain score distribution in the PRO-ACT dataset | b) Bulbar subdomain score distribution in the ALS TDI dataset |
| 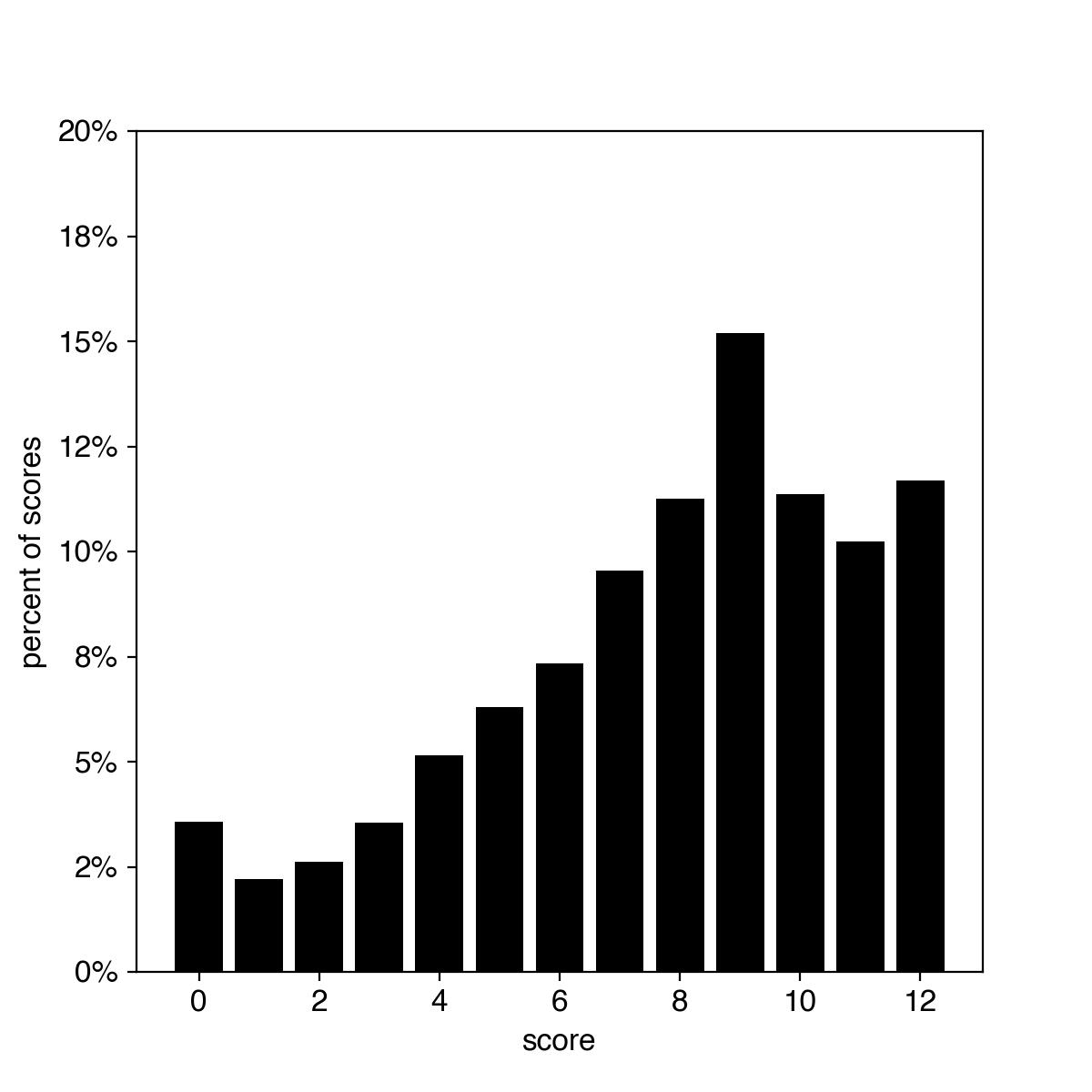 | 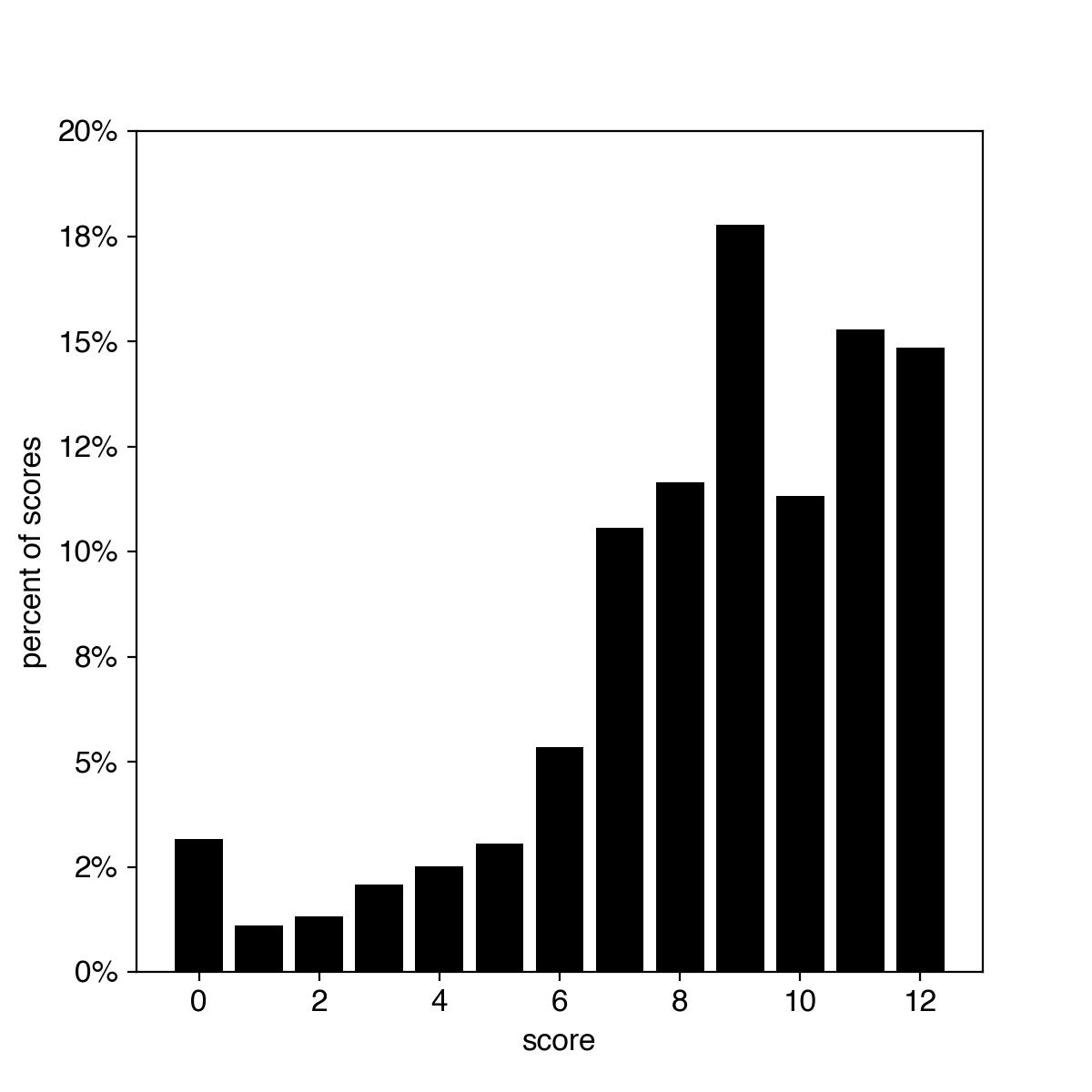 |
| c) Fine Motor subdomain score distribution in the PRO-ACT dataset | d) Fine Motor subdomain score distribution in the ALS TDI dataset |
| 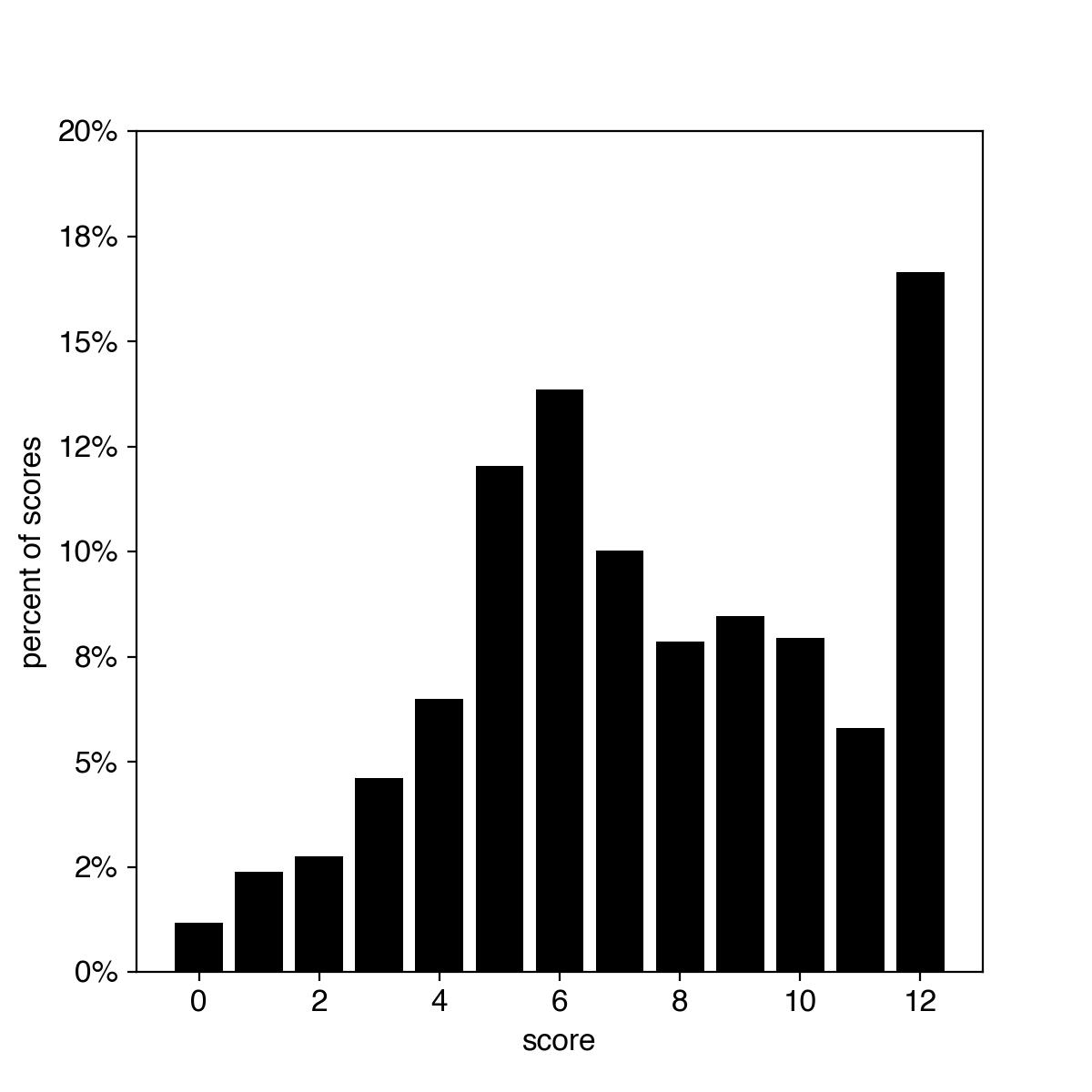 | 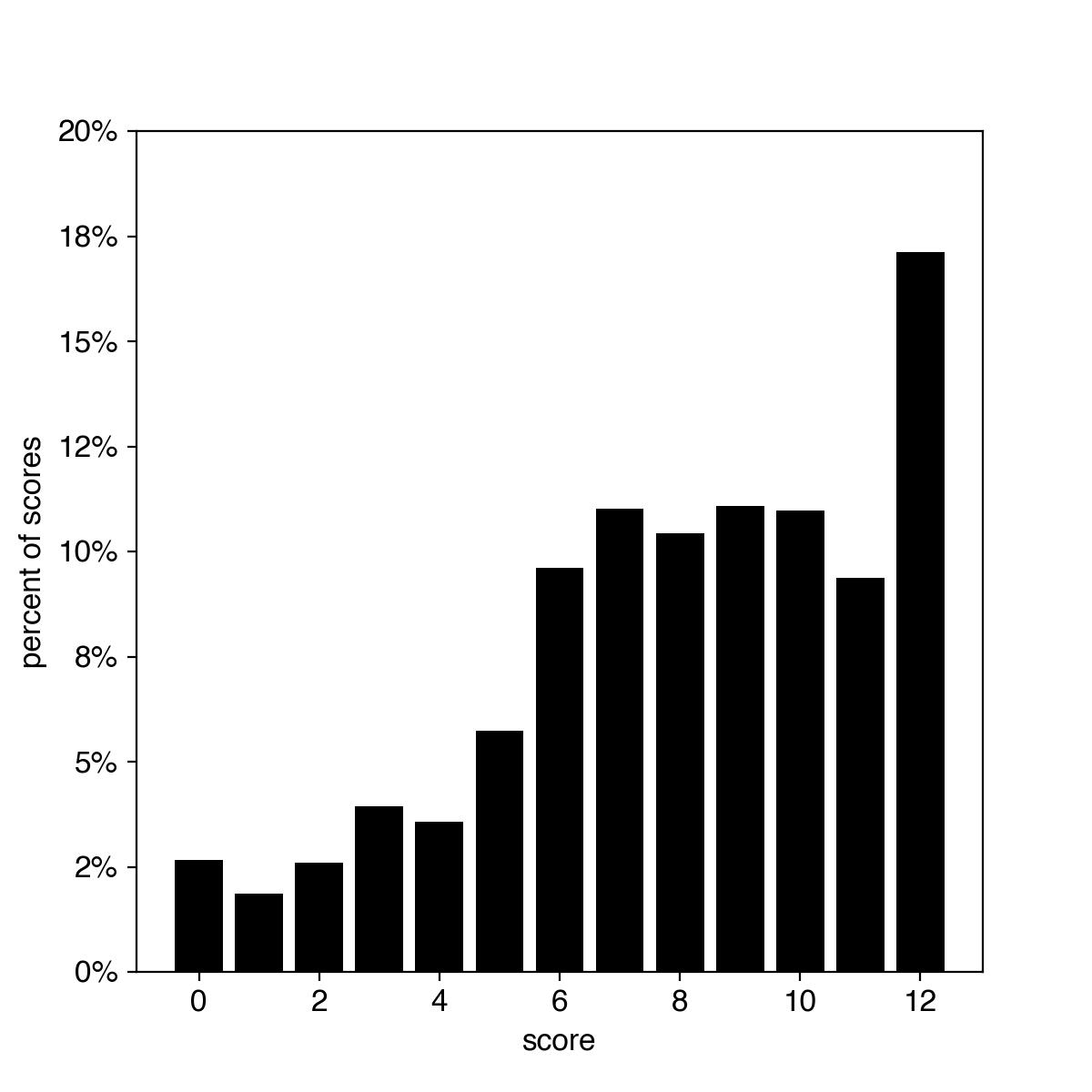 |
| e) Gross Motor subdomain score distribution in the PRO-ACT dataset | f) Gross Motor subdomain score distribution in the ALS TDI dataset |
| 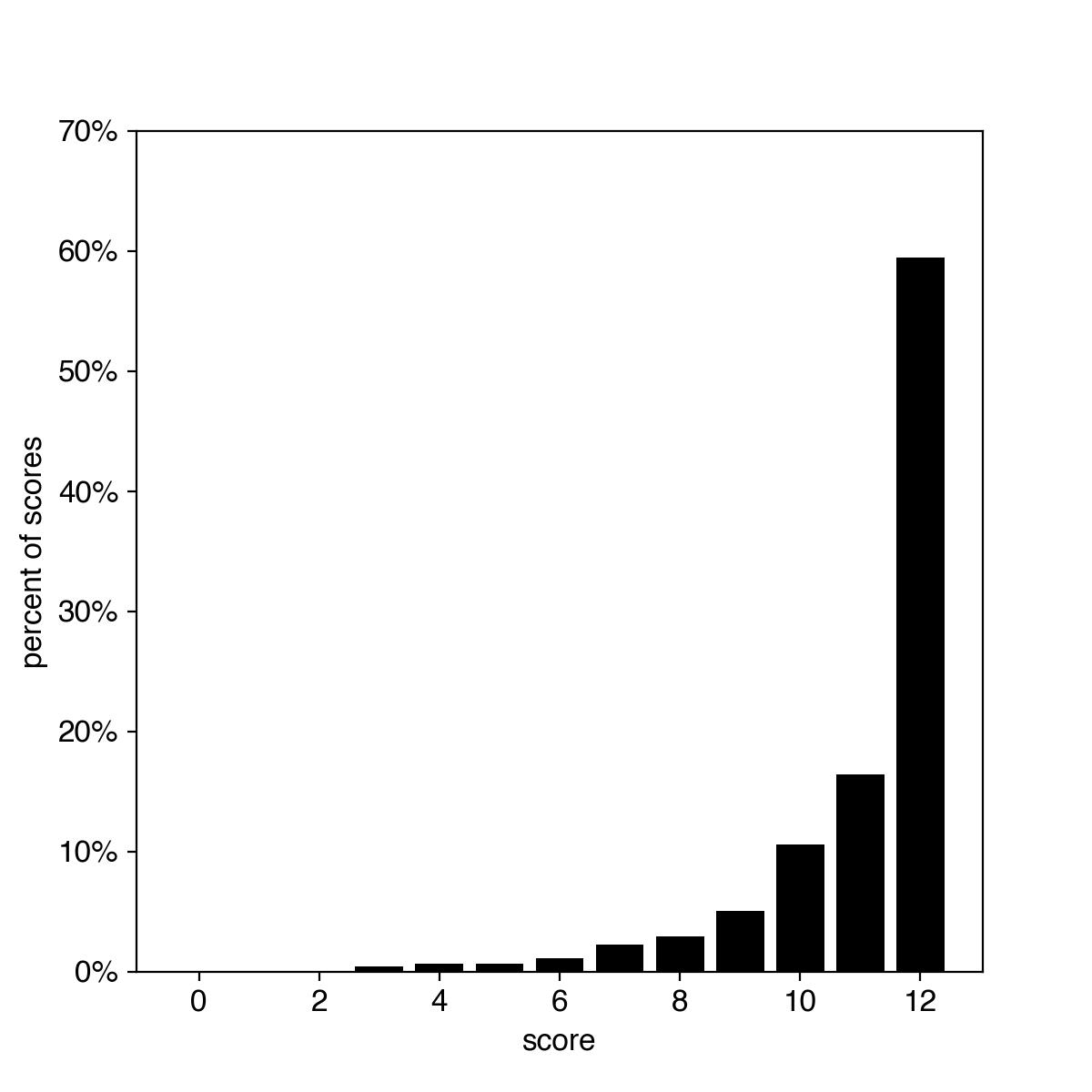 | 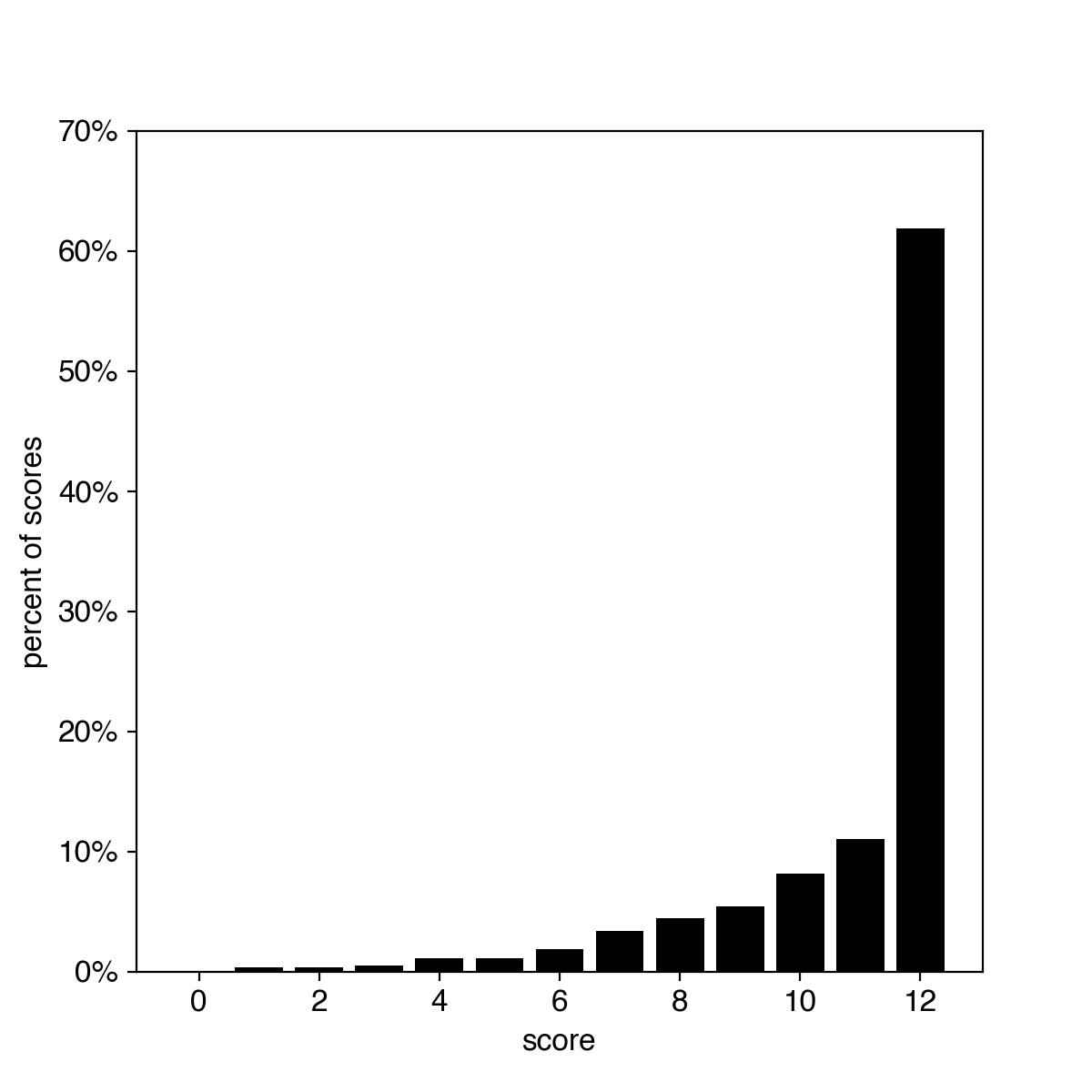 |
| g) Respiratory subdomain score distribution in the PRO-ACT dataset | h) Respiratory subdomain score distribution in the ALS TDI dataset |

**Supplemental Figure 4. ALSFRS-R subdomain score distribution from baseline to 24 weeks in the PRO-ACT (n=792) and ALS TDI (n=1,108) datasets**. Subdomains comprise 3 items scored from 4 (normal - no symptoms) to 0 (severe symptoms) and sum to a score between 12 (normal - no symptoms) and 0 (severe symptoms). Percent of score is defined as count of score divided by total count of all scores. Maximum value of y-axis varies between subdomains to aid comparison between datasets.

**Supplemental Table 2. ALSFRS-R subdomain and total score mean (sd) [confidence interval] actual and percent change at 24 weeks in the PRO-ACT (n=792) and ALS TDI (n=1,108) datasets**.

| **Subdomain** | **PRO-ACT** | | **ALS TDI** | |
| --- | --- | --- | --- | --- |
|  | **actual change** | **% change** | **actual change** | **% change** |
| Total Score | -5.6 (0.1)  [-6.5 \| -4.8] | -11.7% (0.2%)  [-13.5% \| -10.0%] | -3.0 (0.2)  [-3.5 \| -2.6] | -6.3% (0.4%)  [-7.2% \| -5.4%] |
| Bulbar | -1.2 (0.1)  [-1.4 \| -0.9] | -9.6% (0.4%)  [-11.9% \| -7.3%] | -0.6 (0.1)  [-0.7 \| -0.4] | -4.6% (0.5%)  [-5.8% \| -3.3%] |
| Fine Motor | -1.9 (0.1)  [-2.2 \| -1.5] | -15.7% (0.8%)  [-18.6% \| -12.8%] | -1.0 (0.1)  [-1.2 \| -0.9] | -8.4% (0.8%)  [-9.7% \| -7.1%] |
| Gross Motor | -1.7 (0.0)  [-2.0 \| -1.4] | -14.2% (0.3%)  [-16.7% \| -11.7%] | -1.0 (0.0)  [-1.1 \| -0.9] | -8.4% (0.4%)  [-9.7% \| -7.1%) |
| Respiratory | -0.9 (0.1)  [-1.2 \| -0.6] | -7.5% (0.5%)  [-10.1% \| -4.9%] | -0.5 (0.1)  [-0.6 \| -0.3] | -3.8% (0.5%)  [-5.1% \| -2.6%] |

| 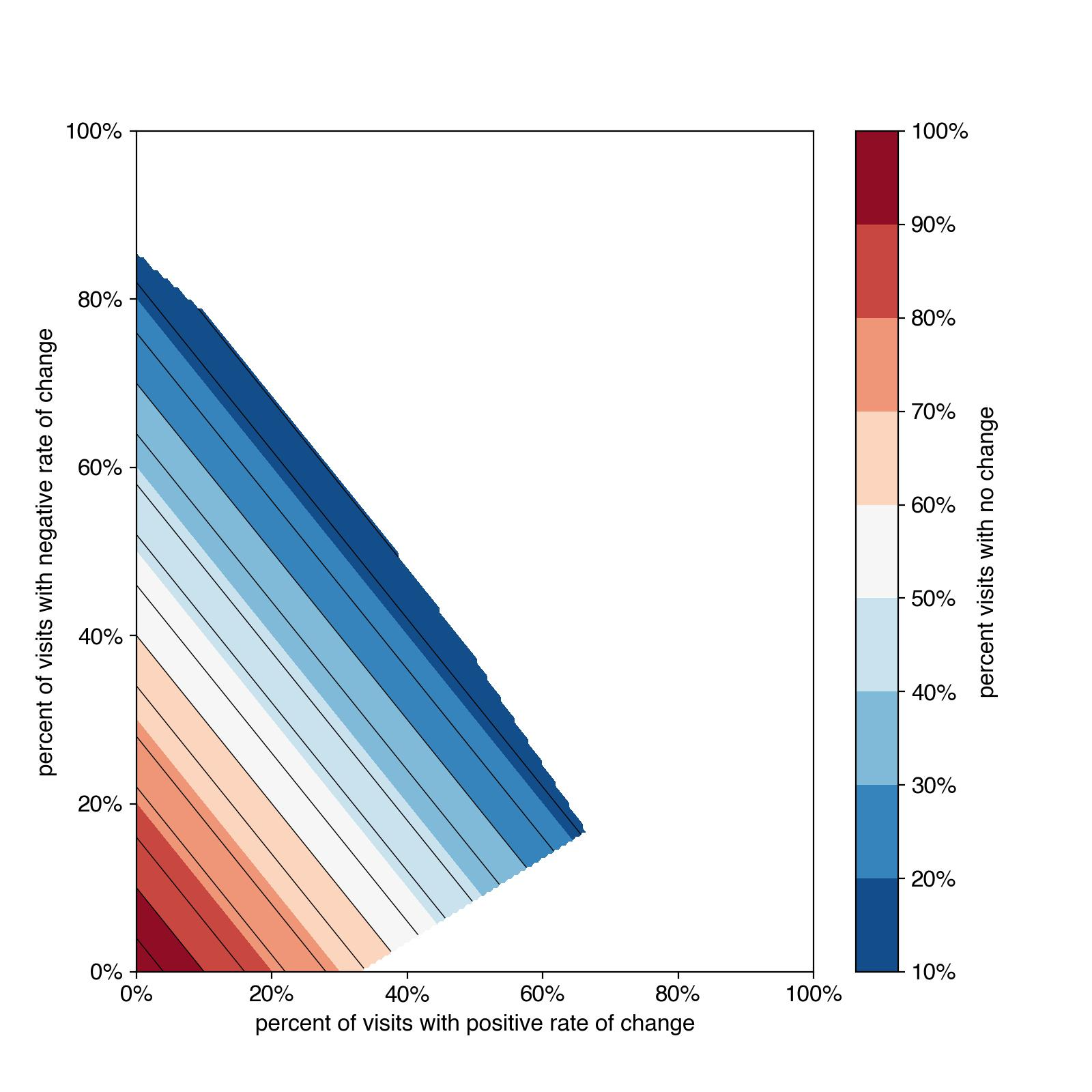 | 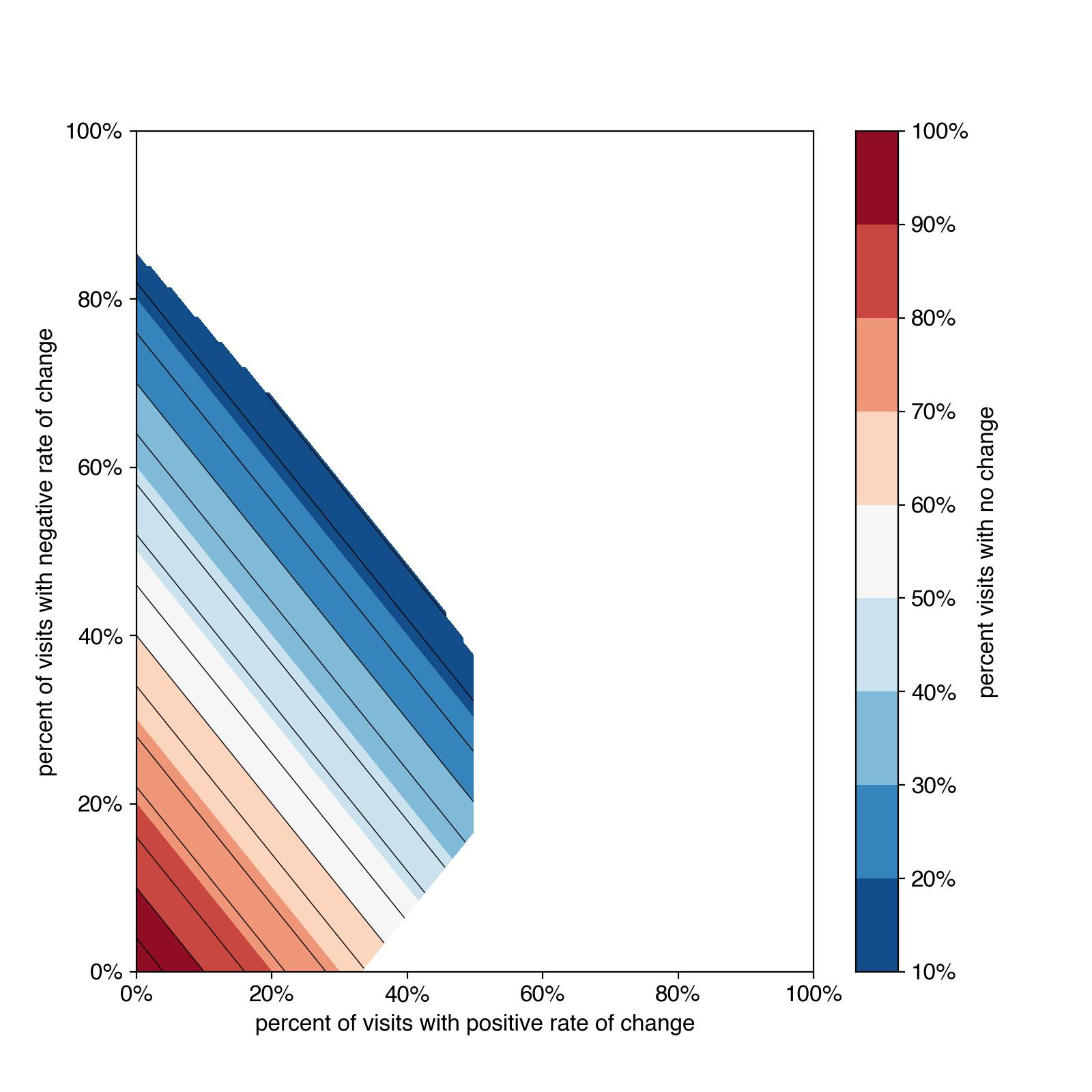 |
| --- | --- |
| a) PRO-ACT patient percent of time on study (N of subjects with > 5 visits is 608). | b) ALS TDI patient percent of time on study (N of subjects with > 5 visits is 479) |

**Supplemental Figure 5. Contour plots for a) PRO-ACT and b) ALS TDI of percent of time on study in negative, no and positive change (slope) in ALSFRS-R total score in subjects with 5 or more visits.** Axes are defined as follows: positive change (x-axis), negative change (y-axis) and no change (z-axis).

| 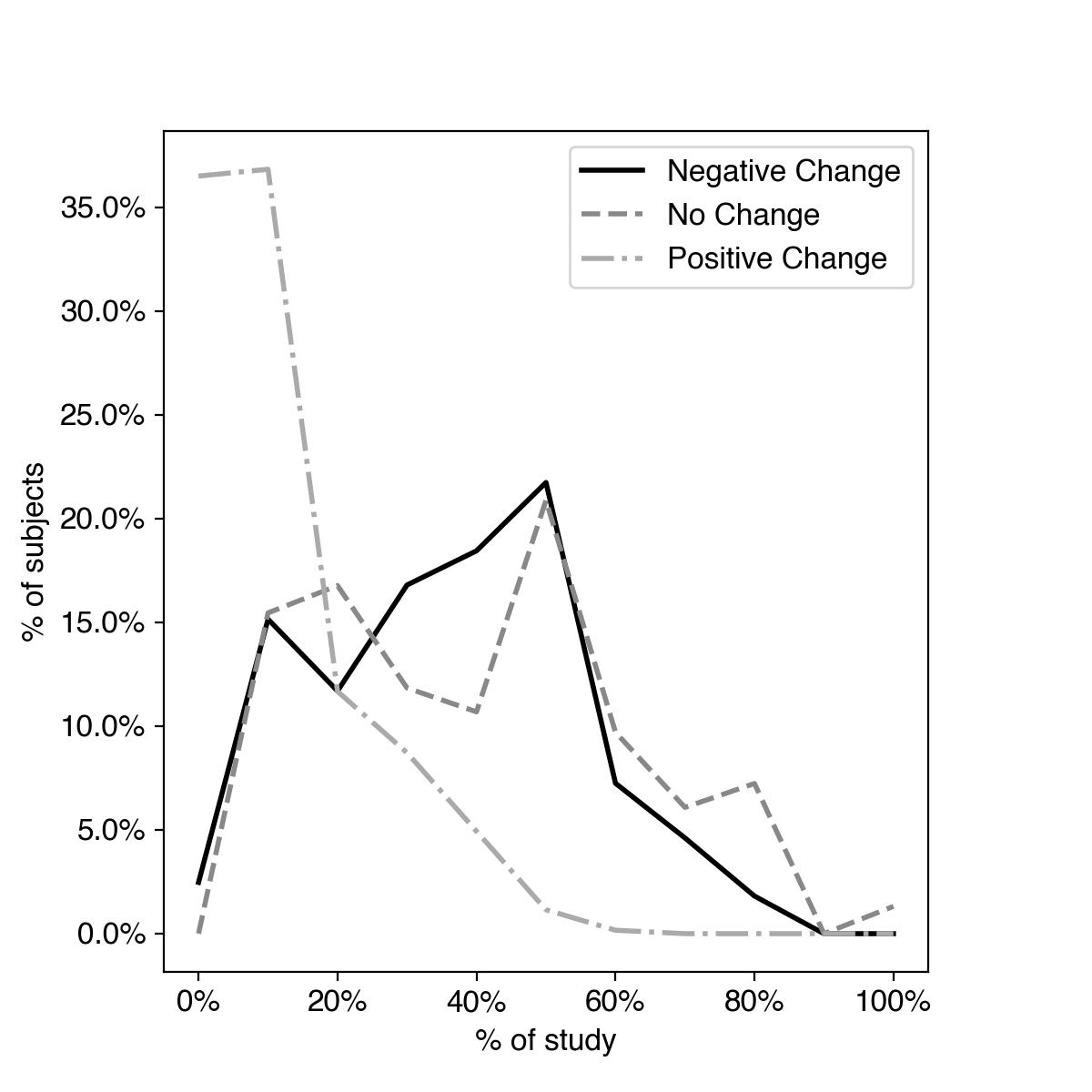 | 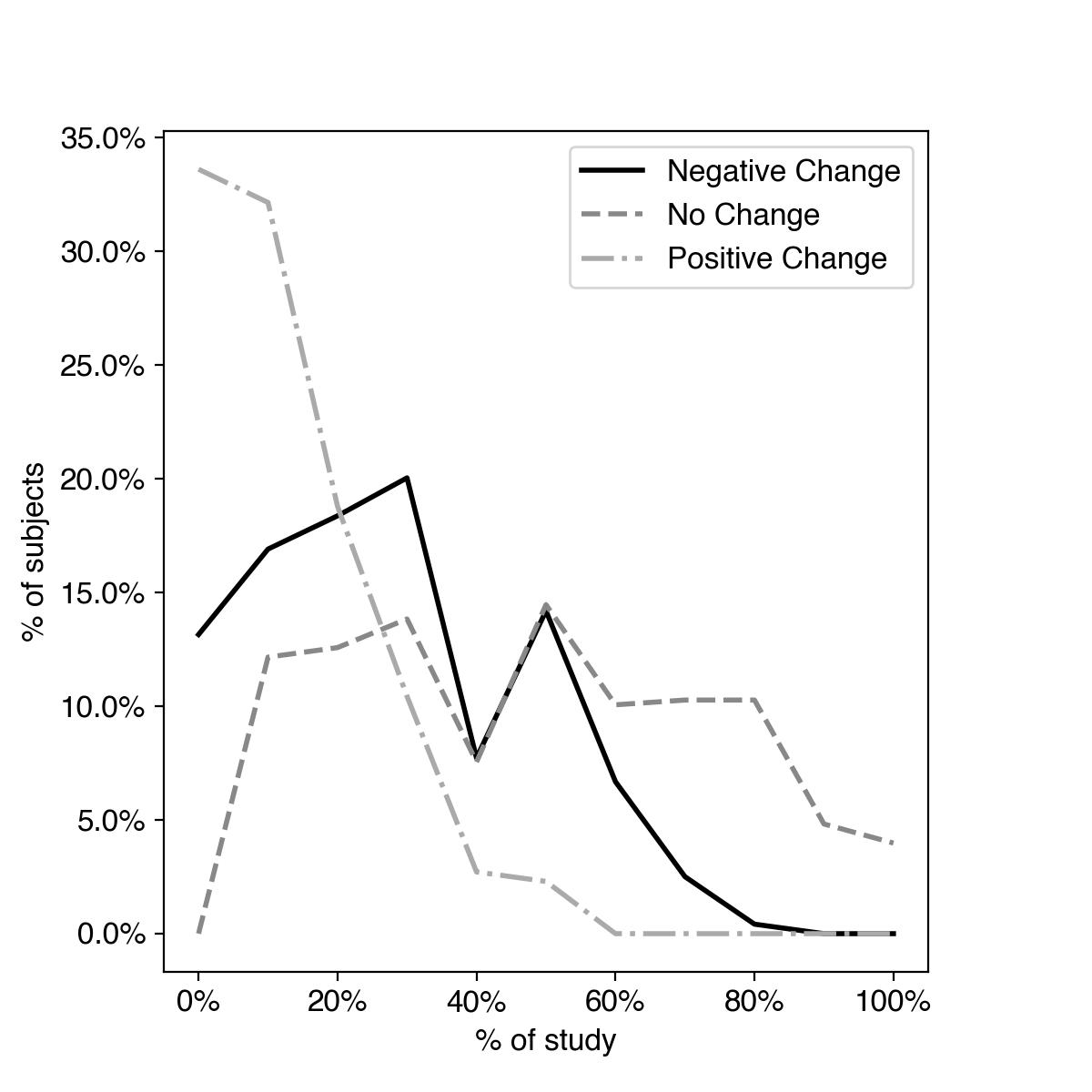 |
| --- | --- |
| a) PRO-ACT percent of subjects in each progression change category by percent of time on study with > 5 visits (N=608) | b) ALS TDI percent of subjects in each progression change category by percent of time on study with > 5 visits (N=479) |

**Supplemental Figure 6.** Percent of subjects in each progression change category by percent of time on study for a) PRO-ACT and b) ALS TDI datasets. Subject total time on study is limited to the first 24 weeks and subjects must have 5 or more visits within this timeframe.

**Supplemental Table 3. Number of subjects stratified as fast or slow by the non-linear and linear progression classification models in the mock “retrospective” data from weeks 0 to 24 in the ALS TDI dataset.**  For the non-linear model the definition of fast progressors in Table 2 was used. For the linear model 2 slope definitions were used: -0.9 points/month[^21^](https://docs.google.com/document/d/13V8ijBiGLOWaytl_Pp8N78dzGi-6gKRS/edit#bookmark=id.bvwbub2mb3ht) and then by -1.5 points/month[^19^](https://docs.google.com/document/d/13V8ijBiGLOWaytl_Pp8N78dzGi-6gKRS/edit#bookmark=id.6wvhsnxdk3om) change in ALSFRS-R total score. A total of 548 subjects had the required 5 or more visits in the mock “retrospective” data. Table cells show count of subjects by cohort and model with percent of total subjects in parentheses.

| **Cohort** | **Linear model**  **(-0.9 points/month)** | **Linear model**  **(-1.5 points/month)** | **Non-linear Model**  **(> 50% of time with negative decline)** |
| --- | --- | --- | --- |
| **Fast** | **143 (26%)** | **65 (12%)** | **92 (17%)** |
| **Slow** | **405 (74%)** | **483 (88%)** | **456 (83%)** |
| **Total** | **548** | **548** | **548** |

**Supplemental Table 4. Mean decline [confidence interval] in 14 day rolling subdomain scores between week minus 24 and zero (retrospective data) and week zero and 24 (prospective data).** Of 548 eligible subjects in the retrospective dataset, 488 subjects have data to week 24 in the prospective study (week 48 in the ALS TDI dataset). Table 4a tabulates results for the non-linear model, 4b for the linear model with fast progressors slope threshold of -0.9 points/month, and 4c for the linear model with fast progressor slope threshold of -1.5 points/month.

| ***ALSFRS-R subdomain*** | **Fast Progressors** | | **Slow Progressors** | |
| --- | --- | --- | --- | --- |
|  | **Retrospective (n=548)** | **Prospective**  **(n=488)** | **Retrospective (n=548)** | **Prospective**  **(n=488)** |
| Total score | -7.5 [-13.3,-2.4] | -6.8 [-12.0,-2.6] | -2.5 [-7.0,0.5] | -2.2 [-9.2,2.0] |
| Bulbar subdomain score | -1.4 [-3.3,0.0] | -1.0 [-3.4,0.0] | -0.5 [-2.0,0.0] | -0.2 [-2.0,1.0] |
| Fine motor subdomain score | -2.8 [-5.3,0.0] | -3.2 [-5.6,-1.2] | -0.9 [-3.0,0.5] | -1.1 [-4.0,0.0] |
| Gross motor subdomain score | -2.3 [-5.3,0.6] | -2.2 [-5.4,-0.2] | -0.8 [-3.0,0.0] | -0.8 [-4.2,1.0] |
| Respiratory subdomain score | -0.9 [-3.0,0.0] | -0.4 [-1.6,0.0] | -0.3 [-3.0,1.0] | -0.1 [-2.0,1.0] |

a) Non-linear model with fast progressors threshold in table 2

| ***ALSFRS-R subdomain*** | **Fast Progressors** | | **Slow Progressors** | |
| --- | --- | --- | --- | --- |
|  | **Retrospective (n=548)** | **Prospective**  **(n=488)** | **Retrospective (n=548)** | **Prospective**  **(n=488)** |
| Total score | -8.2 [-15.8,-4.4] | -4.0 [-16.6,2.0] | -1.7 [-5.0,1.2] | -1.9 [-6.0,1.3] |
| Bulbar subdomain score | -1.5 [-3.0,0.0] | -0.2 [-4.3,1.0] | -0.4 [-2.0, 0.0] | -0.3 [-2.0,0.0] |
| Fine motor subdomain score | -2.7 [-7.2,0.0] | -2.1 [-6.0,0.0] | -0.7 [-2.0,1.0] | -0.8 [4.0,0.0] |
| Gross motor subdomain score | -2.6 [-4.6,-1.0] | -1.3 [-5.0,1.0] | -0.5 [-2.0,-1.0] | -0.7 [-2.0,0.7] |
| Respiratory subdomain score | -1.5 [-4.0,0.6] | -0.3 [-3.3,1.0] | -0.1 [-2.0,-1.0] | 0.0 [-1.0,0.7] |

b) Linear model with fast progressors slope threshold -0.9 points/month

| ***ALSFRS-R subdomain*** | **Fast Progressors** | | **Slow Progressors** | |
| --- | --- | --- | --- | --- |
|  | **Retrospective (n=548)** | **Prospective**  **(n=488)** | **Retrospective (n=548)** | **Prospective**  **(n=488)** |
| Total score | -12.1 [-17.0,-9.0] | -10.7 [-18.0,-2.5] | -2.4 [-7.0. 0.2] | -2.0 [-8.0,2.0] |
| Bulbar subdomain score | -2.0 [-3.0,-0.4] | -2.2 [-5.0,-0.3] | -0.5 [-2.0,0.0] | -0.2 [-2.0,1.0] |
| Fine motor subdomain score | -4.9 [-8.0,-3.0] | -3.7 [-5.8,-2.0] | -0.8 [-3.0,-2.8] | -1.0 [-4.0,0.0] |
| Gross motor subdomain score | -3.6 [-5.2,-2.4] | -2.7 [-5.8,0.0] | -0.8 [-3.0,0.2] | -0.8 [-3.0,1.0] |
| Respiratory subdomain score | -1.4 [-3.0,0.2] | -2.2 [-4.0,0.8] | -0.3 [-3.0,1.0] | -0.0 [-1.0,1.0] |

c) Linear model with fast progressors slope threshold -1.5 points/month
